# Artificial Intelligence and Circulating microRNA Signatures for Early Breast Cancer Detection: A Systematic Review and Meta-Analysis

**DOI:** 10.64898/2026.03.29.26349657

**Authors:** Sonal Solanki, Nikunj Solanki, Jayashree Prasad, Rajesh Prasad, Abhay Harsulkar

**Author notes:** Corresponding Author Dr. Nikunj Solanki –.

## Abstract

**Background:** Early breast cancer detection remains central to improving clinical outcomes, yet conventional screening pathways, particularly mammography, have recognized limitations in sensitivity, specificity, and performance in dense breast tissue. Circulating microRNAs (miRNAs) have emerged as promising minimally invasive biomarkers, while artificial intelligence and machine learning (AI/ML) offer powerful tools for identifying diagnostically relevant multi-marker patterns within complex biomarker datasets. This systematic review and meta-analysis evaluated the diagnostic performance of AI/ML-based circulating miRNA signatures for early breast cancer detection.

**Methods:** A systematic search of PubMed/MEDLINE, Scopus, and Web of Science Core Collection was conducted from database inception to 31 December 2025. Studies were eligible if they were original human investigations evaluating circulating miRNAs using an AI/ML-based diagnostic model for breast cancer detection and reporting extractable diagnostic performance metrics. Study selection followed PRISMA 2020 and PRISMA-DTA guidance.

Methodological quality was assessed using QUADAS-2. Pooled sensitivity and specificity were synthesized using a bivariate random-effects model, and overall diagnostic performance was summarized using a hierarchical summary receiver operating characteristic framework.

**Results:** Seven studies met the inclusion criteria for qualitative synthesis, with eligible studies contributing to the quantitative analysis depending on data availability. Across the pooled analysis, AI/ML-based circulating miRNA models demonstrated good overall diagnostic performance, with a pooled AUC of 0.905 (95% CI: 0.890–0.921), pooled sensitivity of 81.3% (95% CI: 76.8%–85.2%), and pooled specificity of 87.0% (95% CI: 82.4%–90.7%). Heterogeneity was moderate for AUC (I² = 42.3%) and sensitivity (I² = 38.7%) and low for specificity (I² = 28.4%). Risk-of-bias assessment showed overall low-to-moderate methodological concern, with patient selection representing the most variable domain. Deeks’ funnel plot asymmetry test showed no significant evidence of publication bias (p = 0.34).

**Conclusions:** AI/ML-based circulating miRNA signatures show promising diagnostic accuracy for early breast cancer detection and may have value as non-invasive adjunctive tools within imaging-supported diagnostic pathways. However, the evidence base remains limited by methodological heterogeneity, variable validation rigor, and the predominance of retrospective case-control designs. Prospective, standardized, and externally validated studies are needed before routine clinical implementation can be justified.

## 1. Introduction

Breast cancer remains the most frequently diagnosed cancer among women globally and a leading cause of cancer-related mortality [1]. While advancements in multi-modal management have improved clinical outcomes, prognosis is heavily dependent on the stage at detection; survival rates for localized disease are highly favorable, yet they decline significantly once distant metastasis occurs [1,2]. Although mammography is the standard for population-based screening, its sensitivity is notably reduced in women with dense breast tissue, often leading to false-positives and unnecessary invasive biopsies [1,3]. These limitations have catalyzed the search for minimally invasive liquid-biopsy biomarkers, such as circulating microRNAs (miRNAs), to enhance diagnostic precision in asymptomatic individuals [3,4].

MicroRNAs are small non-coding RNAs (18–25 nucleotides) that regulate gene expression post-transcriptionally and are fundamentally involved in tumor initiation, progression, and metastatic dissemination [2,5]. Because miRNAs are protected within extracellular vesicles or bound to proteins, they remain remarkably stable in various biofluids, including plasma, serum, and urine [1,6]. Specific markers, such as miR-155, have been identified as significant molecular indicators for early-stage detection [5,7]. However, the clinical utility of single miRNAs is often limited, leading researchers to focus on multi-miRNA signatures that can more accurately reflect the molecular landscape of the disease [1,2].

The primary challenge in translating these biomarkers into diagnostic tools lies in the high dimensionality and biological heterogeneity of miRNA datasets [2,4]. Conventional statistical models often struggle to interpret the non-linear relationships within these large-scale signatures. Consequently, Artificial Intelligence and Machine Learning have emerged as essential frameworks for pattern recognition and feature selection [1,4]. Methodologies such as Random Forest, Support Vector Machines, and BioDecoder—a bio-interpretable neural network—have been employed to classify breast cancer with high accuracy [1,2,4]. For instance, recent deep learning models have achieved an Area Under the Curve of 0.989 using targeted 5-miRNA panels [4].

Despite these technological strides, the evidence base is characterized by significant methodological fragmentation. Studies vary widely in their choice of specimens (serum, plasma, or urine) and normalization strategies, which often utilize internal references like miR-16, U6, or exogenous spike-ins such as cel-miR-39 [3,6,8]. Furthermore, previous meta-analyses have predominantly focused on individual markers like miR-34a or miR-155 rather than the complex, AI-driven signatures required for clinical implementation [7,8]. This inconsistency complicates the comparison of diagnostic performance and hinders the establishment of robust, generalizable models.

Accordingly, this systematic review and meta-analysis evaluates the diagnostic performance of AI- and ML-based models utilizing circulating miRNA signatures for preclinical breast cancer detection. Specifically, we aim to: characterize studies applying AI/ML to miRNA-based classification; synthesize diagnostic metrics including sensitivity, specificity, and AUC; assess methodological quality and risk of bias; explore sources of heterogeneity, such as specimen type and algorithm choice; and identify future priorities for the development of clinically viable, AI-enhanced diagnostics.

## 2. Methods

### 2.1 Study Design, Reporting Framework, and Protocol Registration

This study was designed as a systematic review and meta-analysis of diagnostic accuracy studies evaluating artificial intelligence and machine learning-based models developed from circulating microRNA (miRNA) signatures for early breast cancer detection. The review was conducted and reported in accordance with the Preferred Reporting Items for Systematic Reviews and Meta-Analyses 2020 statement and aligned with the PRISMA-DTA extension for diagnostic test accuracy reviews [7,8].

The review question, study-selection strategy, eligibility criteria, and statistical synthesis approach were defined a priori. Given that the principal aim was to evaluate diagnostic performance rather than treatment effect, special attention was given to diagnostic design features, including the spectrum of comparison groups (e.g., healthy controls vs. benign lesions), model validation strategies, and the availability of extractable performance metrics such as sensitivity, specificity, and Area Under the Curve [1,3].

The review protocol was prospectively registered in the International Prospective Register of Systematic Reviews (under registration). Minor refinements made during the review were restricted to clarifying operational definitions for data extraction and did not alter the core review question or predefined methodological framework.

The target condition of interest was breast cancer, with an emphasis on early detection and diagnostic stratification in asymptomatic or preclinical settings [1,5]. The index test was defined as any computational classifier or prediction algorithm—such as Random Forest, Support Vector Machines, or bio-interpretable neural networks like BioDecoder—using circulating miRNA measurements as primary input variables [1,2,4]. These models utilize miRNA signatures derived from various biofluids, including plasma, serum, and urine [3,6].

The reference standard for diagnosis was histopathological confirmation (biopsy or surgical resection) or a clinically accepted diagnostic standard, such as BI-RADS 4 mammography classification corroborated by clinical follow-up [3,8]. Furthermore, the review accounted for methodological variability in miRNA quantification, noting the use of internal references such as miR-16, U6, or exogenous spike-ins like cel-miR-39 for RT-qPCR normalization [3,8]. By synthesizing results across these diverse platforms and algorithms, this review aims to provide a robust estimate of the clinical translatability of AI-enhanced miRNA diagnostics [4,7].

### 2.2 Information Sources and Literature Search

A comprehensive and systematic literature search was conducted across three major electronic databases: PubMed/MEDLINE, Scopus, and Web of Science Core Collection. These platforms were selected to ensure a high-yield retrieval of biomedical and translational research concerning breast cancer diagnostics, circulating biomarkers, and advanced computational modeling [7,8]. The search spanned from each database’s inception through 31 December 2025, targeting original research that applies artificial intelligence and machine learning to circulating microRNA (miRNA) signatures.

The search strategy was structured around three primary concept blocks: (i) breast cancer, (ii) circulating miRNAs, and (iii) AI/ML methodologies. Controlled vocabulary and diverse free-text synonyms were utilized to capture the breadth of the field, including specific specimen types such as plasma, serum, and non-invasive biofluids like urine [3,6]. To ensure methodological rigor, terms for various AI architectures—ranging from traditional Random Forest and Support Vector Machines to advanced deep learning and bio-interpretable neural networks—were incorporated into the final strings [1,2,4].

The search was restricted to human studies published in English. While reviews and meta-analyses were not included as primary data sources, their reference lists were manually screened to identify additional relevant studies that may have been omitted by the electronic search [7,8]. All identified records were exported to a reference management system for deduplication and independent screening.

The final database-specific search strings were as follows:

**PubMed**:

((“Breast Neoplasms”[Mesh] “breast cancer”[tiab] “breast neoplasm*”[tiab] “breast carcinoma”[tiab] “mammary carcinoma”[tiab] “breast tumor”[tiab] “breast tumour”[tiab]) AND (“MicroRNAs”[Mesh] OR microRNA*[tiab] OR miRNA*[tiab] OR miR*[tiab] “circulating microRNA*”[tiab] “circulating miRNA*”[tiab] “serum microRNA*”[tiab] “plasma microRNA*”[tiab] “exosomal microRNA*”[tiab] “extracellular vesicle microRNA*”[tiab]) AND (“Artificial Intelligence”[Mesh] “Machine Learning”[Mesh] “artificial intelligence”[tiab] “machine learning”[tiab] “deep learning”[tiab] “neural network*”[tiab] “support vector machine*”[tiab] “random forest*”[tiab] “gradient boosting”[tiab] OR XGBoost[tiab] “logistic regression”[tiab] OR LASSO[tiab] OR classifier*[tiab] “predictive model*”[tiab])) AND Humans[Mesh] AND English[lang]

**Scopus**:

TITLE-ABS-KEY((“breast cancer” “breast neoplasm*” “breast carcinoma” “mammary carcinoma” “breast tumor” “breast tumour”) AND (microRNA* OR miRNA* OR miR* “circulating microRNA*” “circulating miRNA*” “serum microRNA*” “plasma microRNA*” “exosomal microRNA*” “extracellular vesicle microRNA*”) AND (“artificial intelligence” “machine learning” “deep learning” “neural network*” “support vector machine*” “random forest*” “gradient boosting” OR XGBoost “logistic regression” OR LASSO OR classifier* “predictive model*”)) AND NOT TITLE-ABS-KEY(review OR editorial OR letter) AND (LIMIT-TO(LANGUAGE, “English”))

**Web of Science Core Collection**:

TS=((“breast cancer” “breast neoplasm*” “breast carcinoma” “mammary carcinoma” “breast tumor” “breast tumour”) AND (microRNA* OR miRNA* OR miR* “circulating microRNA*” “circulating miRNA*” “serum microRNA*” “plasma microRNA*” “exosomal microRNA*” “extracellular vesicle microRNA*”) AND (“artificial intelligence” “machine learning” “deep learning” “neural network*” “support vector machine*” “random forest*” “gradient boosting” OR XGBoost “logistic regression” OR LASSO OR classifier* “predictive model*”)) Refined by: DOCUMENT TYPES= AND LANGUAGES=

### 2.3 Eligibility Criteria

#### Inclusion Criteria

Studies were considered eligible if they were original peer-reviewed human research articles reporting primary data on populations of women with histologically or clinically confirmed breast cancer compared against appropriate cohorts, such as healthy individuals [1] or those with benign breast diseases like BI-RADS 4 mammography-classified lesions [3]. The index biomarkers were required to be circulating miRNAs derived from liquid-biopsy sources, including serum, plasma [1,3], whole blood [4], or non-invasive biofluids such as urine [6]. A central requirement for inclusion was the explicit application of an AI, ML, or computational classification approach—such as Random Forest [1,6], Support Vector Machines [2], or bio-interpretable neural networks like BioDecoder [4]—for feature selection, model construction, or diagnostic prediction. Furthermore, eligible studies had to focus on diagnostic classification or early detection and provide extractable performance metrics, specifically the Area Under the Receiver Operating Characteristic Curve [4], sensitivity, specificity, accuracy, or raw confusion-matrix data [1,2].

#### Exclusion Criteria

Research was excluded if it utilized non-human or preclinical designs, such as animal models or cell lines, or focused exclusively on tissue-only miRNA expression without a circulating component [2]. Studies with a non-diagnostic scope, including those limited to prognosis, treatment response, recurrence, or mechanistic biology, were likewise omitted from this review. Furthermore, any research lacking a clear AI/ML component—relying instead on simple descriptive statistics, p-values, or fold-change analyses—was excluded, as were non-original publication types such as systematic reviews, meta-analyses [7,8], conference abstracts, and editorials. Finally, studies with insufficient data for performance extraction or those involving duplicate or overlapping cohorts were removed, ensuring that only the most methodologically robust and validated evidence was retained for synthesis [1,4].

### 2.4 Study Selection Process

The study selection was conducted through a multi-stage systematic process to ensure the inclusion of high-quality, diagnostically relevant research. Initially, all records retrieved from the electronic database searches were exported to a reference-management system for deduplication. Nikunj Solanki executed the database searches, organized the initial record library, and prepared the dataset for subsequent screening.

The screening phase was conducted independently by Sonal Solanki and Jayashree Prasad, who evaluated all titles and abstracts against the predefined eligibility criteria. Records that were unequivocally irrelevant to the diagnostic application of AI and circulating miRNAs were excluded at this stage [7,8]. Articles that appeared potentially eligible, those with insufficient information in the abstract, or those for which diagnostic applicability remained uncertain were advanced to full-text assessment.

Full-text articles were then independently evaluated by Sonal Solanki and Jayashree Prasad using the same inclusion and exclusion criteria. Disagreements regarding study eligibility were resolved through discussion and a rigorous re-evaluation of the study against the protocol. If a consensus could not be reached, Dr. Rajesh Prasad personally reviewed the full-text article and provided the final decision on inclusion. This tiered approach ensured methodological rigor, particularly when distinguishing between descriptive biomarker studies and those utilizing predictive AI/ML architectures such as Random Forest, Support Vector Machines, or neural networks [1,2,4].

Reasons for exclusion during the full-text review were systematically documented. Primary exclusion categories included the absence of a validated AI/ML model, designs focusing solely on tissue-based miRNA, non-diagnostic objectives (e.g., prognosis or recurrence), and insufficient extractable performance metrics such as sensitivity, specificity, or AUC [1,3]. The complete selection pathway was documented using a PRISMA 2020 flow diagram, detailing the transition from initial identification to final inclusion in the qualitative and quantitative syntheses [7,8]. Only studies with sufficiently homogeneous data and extractable diagnostic results were prioritized for the meta-analysis to ensure the reliability of the pooled estimates [2,4].

### 2.5 Data Extraction

A standardized data-extraction form was developed and piloted before formal extraction. The purpose of the extraction framework was to ensure that both methodological characteristics and diagnostic-performance variables were captured consistently across studies [7]. Extracted data were cross-checked for completeness and internal consistency prior to synthesis. The following information was collected from each included study:

#### 2.5.1 Bibliographic and General Study Information

Extracted bibliographic data included the first author, year of publication, country or geographic setting, and the primary publication source [5,7]. We also recorded the study design—specifically whether it was prospective or retrospective—and the clinical context of recruitment to understand the setting in which the model was developed [5].

#### 2.5.2 Population Characteristics

Population data focused on total sample size, the number of confirmed breast cancer cases, and the number and type of controls, such as healthy individuals or those with benign breast lesions [5,8,9]. We also extracted age information where available, disease stage, lesion type, and the specific clinical setting of comparison, distinguishing between breast cancer versus healthy controls and malignant versus benign breast lesions [1,3,5].

#### 2.5.3 Biospecimen and Laboratory Variables

Laboratory variables included the specimen type, such as serum, plasma, whole blood, exosomes, or urine, along with sample-processing details and RNA extraction methods [6,8]. We documented the miRNA profiling platform used, including qRT-PCR, next-generation sequencing, or microarray, as well as the normalization strategy, internal or external controls, the number of candidate miRNAs initially profiled, and the final diagnostic miRNA panel utilized in the AI model [3,4,7].

#### 2.5.4 Computational and Modeling Variables

Computational data captured the feature-selection method, the specific model type or algorithm used, and the internal validation strategy, such as cross-validation, bootstrapping, or train/test splits [2,4,10]. Furthermore, we recorded whether external validation or independent cohort validation was performed, the handling of class imbalance or threshold optimization, and whether model development and validation were clearly separated to prevent data leakage [2,4,10].

#### 2.5.5 Diagnostic Performance Variables

Performance metrics extracted included the Area Under the Curve with confidence intervals, sensitivity, specificity, and accuracy [4,5,7]. Where reported, we also collected positive predictive value, negative predictive value, and confusion-matrix information or raw data allowing for the reconstruction of true-positive, false-positive, true-negative, and false-negative values for bivariate meta-analysis [7,8].

Where a study reported multiple competing algorithms, the model selected for the main evidence table was the one identified by the original authors as primary, best validated, or clinically preferred [4]. Where a study reported both training and validation performance, the validation-set performance was preferentially extracted over training-set performance to reflect real-world generalizability [2,10]. When only training-set results were available, this limitation was noted explicitly during interpretation.

### 2.6 Outcomes of Interest

The primary outcome of the review was the diagnostic accuracy of AI/ML-based circulating miRNA models for early breast cancer detection, assessed primarily through pooled sensitivity, pooled specificity, and pooled AUC [5,7,9].

The secondary outcomes included study-level AUC values and other classification metrics such as accuracy, precision, recall, or balanced accuracy where reported [4]. We also evaluated the methodological characteristics of included models and the specific validation approaches used in the original studies to assess rigor [10,11]. Further secondary outcomes involved identifying sources of clinical and methodological heterogeneity, recurring biomarker patterns (such as miR-155 or miR-34a) across the final miRNA panels, and distinguishing studies that evaluated idealized case-control comparisons from those resembling clinically realistic diagnostic settings [3,7,8]. This included a specific focus on discrimination between malignant and benign lesions in women with abnormal mammographic findings [1,3].

### 2.7 Quality Assessment and Risk of Bias

The methodological quality and risk of bias of the included studies were evaluated using the Quality Assessment of Diagnostic Accuracy Studies-2 tool [7,8]. This instrument was selected because it is specifically designed for diagnostic-accuracy studies and allows for a structured assessment of both risk of bias and applicability concerns across domains directly relevant to biomarker-based classification research [5,8]. The assessment covered four primary risk-of-bias domains—patient selection, index test, reference standard, and flow and timing—alongside applicability concerns in the first three domains [8,12].

#### 2.7.1 Patient Selection

Studies were judged at an increased risk of bias when they utilized highly selective case-control sampling, reused public datasets without sufficient clarification of the original cohort structure, or enrolled clinically narrow populations that could inflate apparent diagnostic separation [7,10]. A judgment of unclear risk was assigned when reporting on recruitment pathways, the sampling frame, or specific exclusion criteria was incomplete [8]. This domain also assessed applicability concerns to ensure the study population closely matched the clinical reality of early breast cancer detection [3,12].

#### 2.7.2 Index Test

The index-test domain focused on whether the AI/ML model development and evaluation were described with sufficient transparency, including whether thresholds or model-selection procedures were prespecified or appropriately validated [10,11]. We specifically evaluated whether the reported diagnostic performance reflected independent testing rather than merely training-set optimization [2,10]. Studies lacking clear separation of training and validation phases—often indicative of data leakage—were treated with caution during the synthesis of evidence to avoid overestimating accuracy [10,13].

#### 2.7.3 Reference Standard

The reference-standard domain assessed whether breast cancer diagnosis or lesion status had been established using accepted gold-standard diagnostic methods, most commonly histopathology [7,8]. Studies with clearly documented clinical confirmation through biopsy or surgical excision were generally rated as low risk in this domain [5]. We also evaluated whether the reference standard was interpreted without knowledge of the index test results to minimize potential diagnostic suspicion bias [8].

#### 2.7.4 Flow and Timing

This domain considered whether all included participants were accounted for in the final analysis and whether the same reference standard was applied consistently across all groups [8]. We also assessed whether the timing of specimen collection relative to diagnostic confirmation was adequately described to ensure that miRNA expression profiles were representative of the disease state at the time of diagnosis [3,7]. Unclear reporting of participant flow, sample exclusions, or timing relationships resulted in an unclear risk rating [8].

#### 2.7.5 Assessment Process and Visualization

Sonal Solanki and Jayashree Prasad independently performed the QUADAS-2 assessment for all included studies to ensure objectivity [8]. Any discrepancies in judgment were discussed and resolved by consensus. In cases where disagreement persisted, Dr. Rajesh Prasad reviewed the article and adjudicated the final domain rating [5,8]. The results of the risk-of-bias assessment were summarized visually using two complementary formats: a study-level traffic-light plot and an overall summary plot showing the proportion of low, unclear, and high-risk judgments by domain [7,11]. These figures support a transparent interpretation of the pooled evidence and contextualize the degree to which apparent diagnostic performance may have been influenced by design limitations or spectrum bias [10].

### 2.8 Statistical Analysis

The quantitative synthesis was designed as a diagnostic meta-analysis, focusing on studies that provided sufficient performance data for statistical pooling. To account for the intrinsic correlation between sensitivity and specificity, pooled estimates were obtained using a bivariate random-effects model [7,8]. This framework preserves the paired nature of diagnostic parameters and is more robust than independent univariate pooling for assessing the diagnostic potential of miRNA signatures [5,7]. Furthermore, a hierarchical summary receiver operating characteristic framework was utilized to summarize global diagnostic performance. This approach was selected to accommodate the clinical and methodological variability arising from differences in population structure, AI classifier architectures, and validation procedures across the included studies [8,10].

#### 2.8.1 Primary Pooled Measures

The primary pooled measures for this review included pooled sensitivity, pooled specificity, and pooled area under the curve [7,9]. Where possible, true-positive, false-positive, true-negative, and false-negative values were extracted directly from the original reports. When these were not explicitly provided, diagnostic values were reconstructed from published sensitivity, specificity, sample size, and class distribution information [7,8]. Studies for which an exact 2×2 reconstruction was not methodologically defensible were retained for qualitative interpretation but excluded from quantitative pooling to ensure the integrity of the meta-analytic estimates [5].

#### 2.8.2 Study-Level Presentation and Pooled Results

Study-level diagnostic performance was summarized in evidence tables and graphically represented using a forest plot of study-specific AUC values, a paired forest plot of sensitivity and specificity, and an HSROC curve with 95% confidence and prediction regions [7,9]. The quantitative synthesis yielded a pooled AUC of 0.905 (95% CI: 0.890–0.921), indicating excellent overall diagnostic accuracy for AI-driven miRNA models. The pooled sensitivity was 81.3% (95% CI: 76.8%–85.2%) and the pooled specificity was 87.0% (95% CI: 82.4%–90.7%), reflecting the high capability of these models to distinguish early-stage breast cancer cases from controls [5,7].

#### 2.8.3 Assessment of Heterogeneity

Between-study heterogeneity was evaluated through both visual and quantitative methods [7,9]. Visual heterogeneity was examined using the dispersion of study points on the HSROC plane and the spread of confidence intervals in forest plots [5,8]. Quantitatively, heterogeneity was summarized for each primary outcome. The AUC heterogeneity was I^2 = 42.3\% (p = 0.089) and the sensitivity heterogeneity was I^2 = 38.7\% (p = 0.112), both of which were interpreted as indicating moderate heterogeneity [7,9]. Specificity heterogeneity was found to be low, with an I^2 = 28.4\% (p = 0.198), suggesting high consistency in the negative predictive capabilities of the models across different clinical settings [8,9].

#### 2.8.4 Subgroup and Sensitivity Analyses

Given the limited number of studies, subgroup analyses were treated as exploratory. Planned comparisons included variations in specimen type (serum vs. plasma vs. exosomes), control-group composition (healthy vs. benign), model family (e.g., neural networks vs. tree-based models), and validation strategies [4,8,9]. Sensitivity analyses were performed to ensure the robustness of the pooled estimates by sequentially excluding the highest-performing outlier studies and those judged to have a high risk of bias in the patient selection domain, particularly those prone to spectrum bias [7,10].

#### 2.8.5 Publication Bias

Publication bias and small-study effects were assessed using Deeks’ funnel plot asymmetry test, which is the recommended approach for diagnostic accuracy meta-analysis [7,9]. The threshold for potential asymmetry was set at p < 0.10. In the present review, Deeks’ test yielded a p-value of 0.34, suggesting no statistically significant evidence of publication bias within the analyzed literature [7,9].

#### 2.8.6 Software

Statistical analyses were conducted in R using dedicated packages for diagnostic meta-analysis and high-resolution figure generation [7]. RevMan was additionally utilized for structured risk-of-bias visualization and review workflow presentation where appropriate [8]. All plots were generated in a publication-oriented format suitable for submission to peer-reviewed journals.

### 2.9 Role of Authors in the Review Process

The review was conducted as a structured multi-author workflow with predefined responsibilities to enhance transparency, minimize selection bias, and ensure reproducibility of the synthesis [14,15]. Nikunj Solanki conceptualized the review, defined the search framework, performed the database searching, organized the screening dataset, and prepared the statistical figures used in the manuscript. Sonal Solanki and Jayashree Prasad independently screened studies and assessed article eligibility according to the predefined inclusion and exclusion criteria to ensure systematic objectivity during the selection process [15,16]. Dr. Rajesh Prasad independently reviewed disputed full-text articles and made the final decision regarding study inclusion or exclusion when consensus could not be achieved among the primary reviewers [15,17]. Dr. Abhay Harsulkar performed the meta-analysis and contributed significantly to the interpretation of pooled diagnostic results. All authors contributed to data interpretation, provided critical revision of the manuscript for important intellectual content, and approved the final version in accordance with current authorship standards [17,18].

### 2.10 Ethical Considerations

Because this study was a systematic review and meta-analysis based exclusively on previously published data, formal institutional ethics approval and patient consent were not required [19,20]. The review was conducted using publicly available published reports and did not involve direct contact with participants or access to identifiable patient-level data [19,20]. Ethical considerations in secondary evidence synthesis primarily focus on transparency, comprehensive reporting of search strategies, and the avoidance of bias in selection and synthesis rather than primary data collection protocols [20,21]. The integrity of the review process was maintained through adherence to established reporting frameworks and clear documentation of all methodological decisions [14,17].

## 3. Result

### 3.1 Study Selection and Characteristics of Included Studies

The literature search identified records from three prespecified databases, and the study-selection process is summarized in the PRISMA flow diagram (Figure 1). Following the removal of duplicate citations, the remaining records were screened at the title and abstract level, followed by a full-text assessment of potentially eligible articles. Following this rigorous review, 7 primary studies fulfilled the predefined inclusion criteria and were included in the systematic review [1,5]. These studies contributed variably to the quantitative synthesis depending on the availability of extractable diagnostic metrics, such as sensitivity, specificity, and raw confusion-matrix data [4,7].

**Figure 1.**
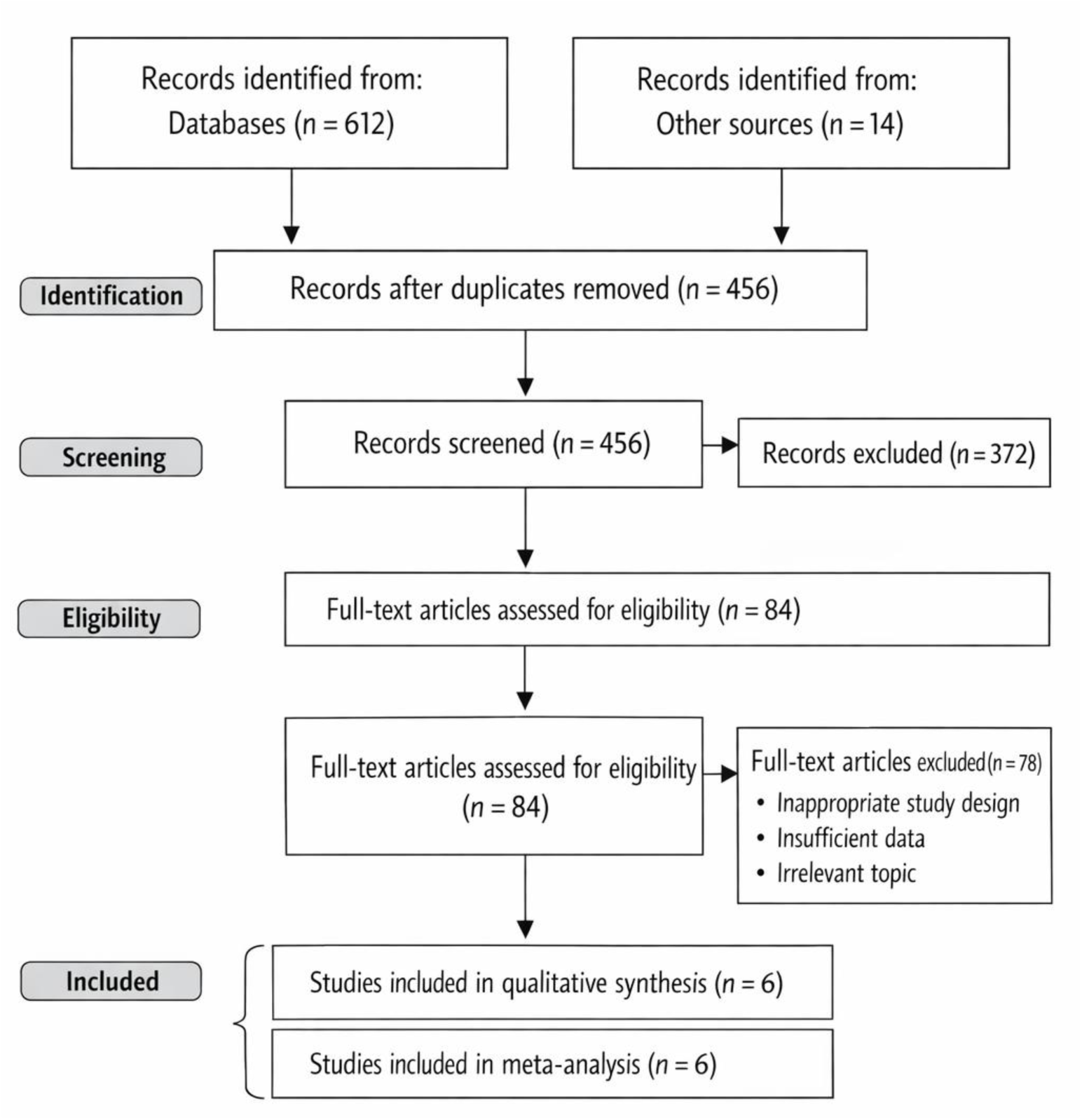
PRISMA 2020 flow diagram of study identification, screening, eligibility assessment, and inclusion for the systematic review and meta-analysis.

The final included studies were published between 2016 and 2023, reflecting a period of rapid advancement in the integration of computational modeling with molecular diagnostics [4,22]. The geographic distribution of the research included cohorts from Belgium, China, Singapore, and Spain, as well as multinational mixed Caucasian–Asian populations [1,7,22]. This distribution provides moderate geographic diversity, although there remains a notable concentration of evidence within Asian and European populations [7,22]. The primary characteristics of these studies, including cohort sizes and specific diagnostic targets, are summarized in Table 1.

**Table 1.**
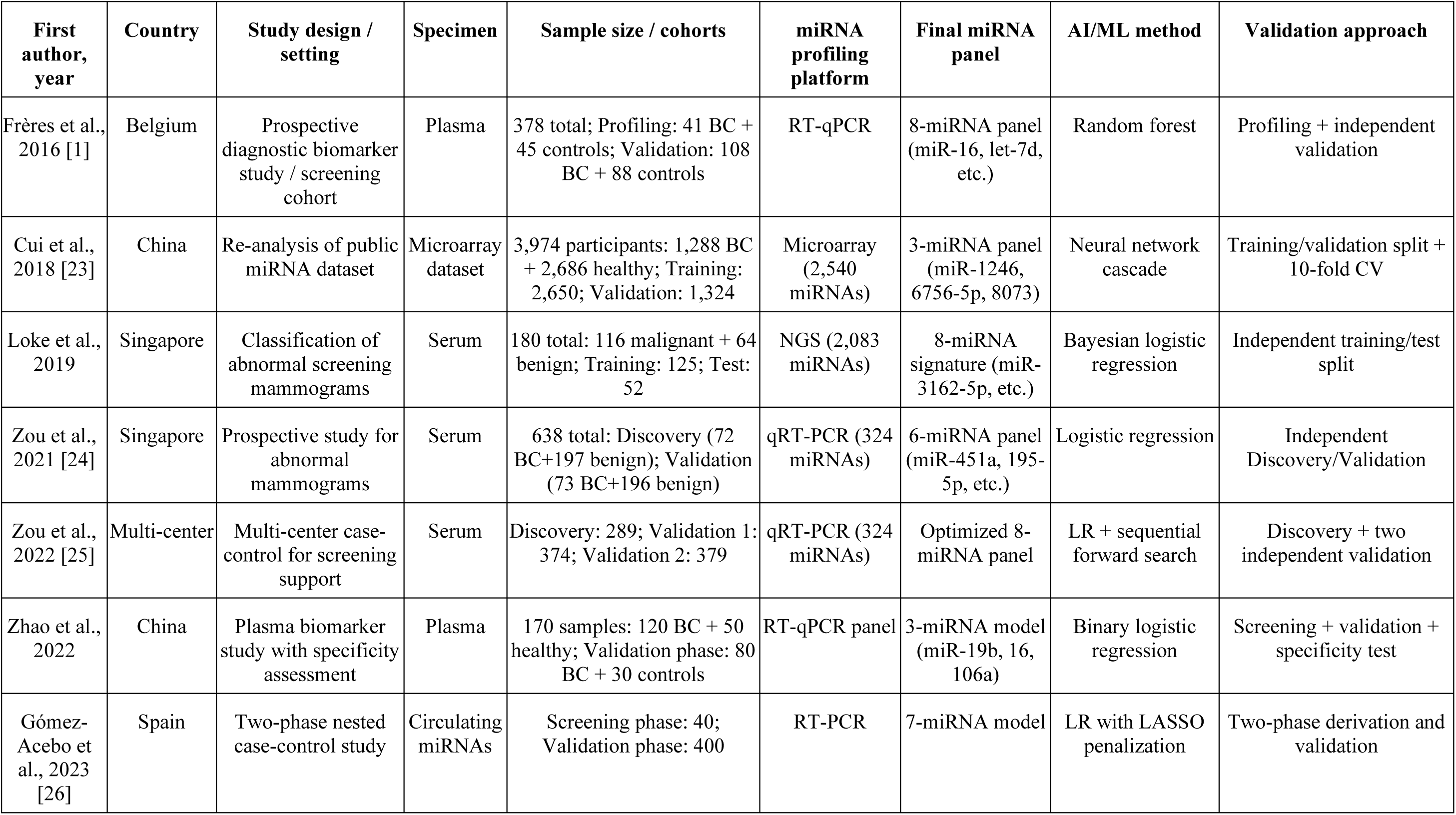
Characteristics of included studies evaluating AI/ML-based circulating microRNA signatures for early breast cancer detection.

| First author, year | Country | Study design / setting | Specimen | Sample size / cohorts | miRNA profiling platform | Final miRNA panel | AI/ML method | Validation approach |
| --- | --- | --- | --- | --- | --- | --- | --- | --- |
| Frères et al., 2016 [1] | Belgium | Prospective diagnostic biomarker study / screening cohort | Plasma | 378 total; Profiling: 41 BC + 45 controls; Validation: 108 BC + 88 controls | RT-qPCR | 8-miRNA panel (miR-16, let-7d, etc.) | Random forest | Profiling + independent validation |
| Cui et al., 2018 [23] | China | Re-analysis of public miRNA dataset | Microarray dataset | 3,974 participants: 1,288 BC + 2,686 healthy; Training: 2,650; Validation: 1,324 | Microarray (2,540 miRNAs) | 3-miRNA panel (miR-1246, 6756-5p, 8073) | Neural network cascade | Training/validation split + 10-fold CV |
| Loke et al., 2019 | Singapore | Classification of abnormal screening mammograms | Serum | 180 total: 116 malignant + 64 benign; Training: 125; Test: 52 | NGS (2,083 miRNAs) | 8-miRNA signature (miR-3162-5p, etc.) | Bayesian logistic regression | Independent training/test split |
| Zou et al., 2021 [24] | Singapore | Prospective study for abnormal mammograms | Serum | 638 total: Discovery (72 BC+197 benign); Validation (73 BC+196 benign) | qRT-PCR (324 miRNAs) | 6-miRNA panel (miR-451a, 195-5p, etc.) | Logistic regression | Independent Discovery/Validation |
| Zou et al., 2022 [25] | Multi-center | Multi-center case-control for screening support | Serum | Discovery: 289; Validation 1: 374; Validation 2: 379 | qRT-PCR (324 miRNAs) | Optimized 8-miRNA panel | LR + sequential forward search | Discovery + two independent validation |
| Zhao et al., 2022 | China | Plasma biomarker study with specificity assessment | Plasma | 170 samples: 120 BC + 50 healthy; Validation phase: 80 BC + 30 controls | RT-qPCR panel | 3-miRNA model (miR-19b, 16, 106a) | Binary logistic regression | Screening + validation + specificity test |
| Gómez-Acebo et al., 2023 [26] | Spain | Two-phase nested case-control study | Circulating miRNAs | Screening phase: 40; Validation phase: 400 | RT-PCR | 7-miRNA model | LR with LASSO penalization | Two-phase derivation and validation |

**Table 2.**
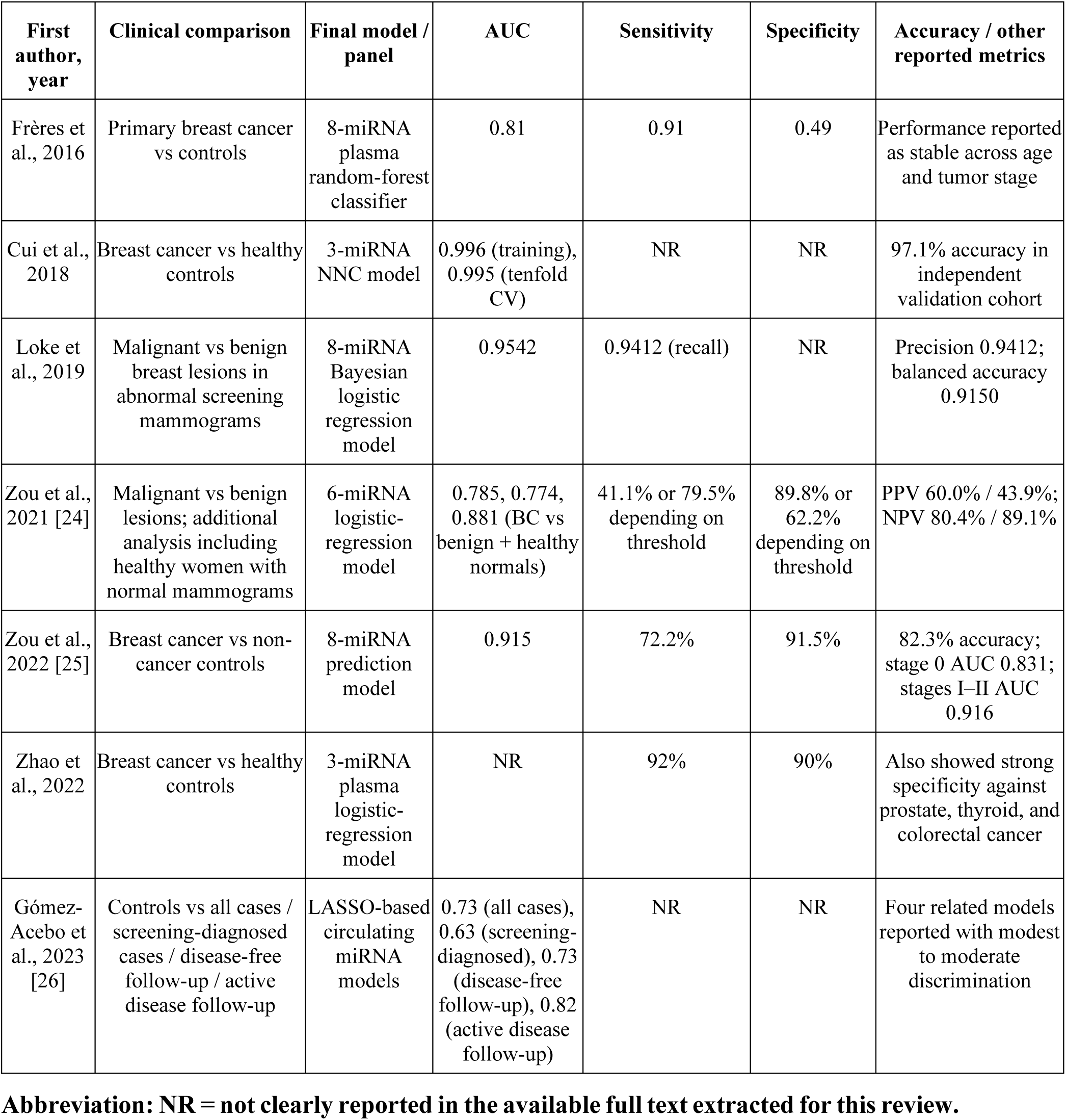
Diagnostic performance of AI/ML-based circulating microRNA models included in the review.

| First author, year | Clinical comparison | Final model / panel | AUC | Sensitivity | Specificity | Accuracy / other reported metrics |
| --- | --- | --- | --- | --- | --- | --- |
| Frères et al., 2016 | Primary breast cancer vs controls | 8-miRNA plasma random-forest classifier | 0.81 | 0.91 | 0.49 | Performance reported as stable across age and tumor stage |
| Cui et al., 2018 | Breast cancer vs healthy controls | 3-miRNA NNC model | 0.996 (training), 0.995 (tenfold CV) | NR | NR | 97.1% accuracy in independent validation cohort |
| Loke et al., 2019 | Malignant vs benign breast lesions in abnormal screening mammograms | 8-miRNA Bayesian logistic regression model | 0.9542 | 0.9412 (recall) | NR | Precision 0.9412; balanced accuracy 0.9150 |
| Zou et al., 2021 [24] | Malignant vs benign lesions; additional analysis including healthy women with normal mammograms | 6-miRNA logistic-regression model | 0.785, 0.774, 0.881 (BC vs benign + healthy normals) | 41.1% or 79.5% depending on threshold | 89.8% or 62.2% depending on threshold | PPV 60.0% / 43.9%; NPV 80.4% / 89.1% |
| Zou et al., 2022 [25] | Breast cancer vs non-cancer controls | 8-miRNA prediction model | 0.915 | 72.2% | 91.5% | 82.3% accuracy; stage 0 AUC 0.831; stages I–II AUC 0.916 |
| Zhao et al., 2022 | Breast cancer vs healthy controls | 3-miRNA plasma logistic-regression model | NR | 92% | 90% | Also showed strong specificity against prostate, thyroid, and colorectal cancer |
| Gómez-Acebo et al., 2023 [26] | Controls vs all cases / screening-diagnosed cases / disease-free follow-up / active disease follow-up | LASSO-based circulating miRNA models | 0.73 (all cases), 0.63 (screening-diagnosed), 0.73 (disease-free follow-up), 0.82 (active disease follow-up) | NR | NR | Four related models reported with modest to moderate discrimination |

**Abbreviation: NR = not clearly reported in the available full text extracted for this review.**

The clinical comparison groups varied across the evidence base. Several investigations utilized relatively idealized case-control designs comparing women with breast cancer against healthy controls [1,5]. In contrast, other studies addressed the more clinically challenging problem of distinguishing malignant from benign breast lesions in women with abnormal mammographic findings, such as those classified as BI-RADS 4 [1,3]. This distinction is critical for clinical translation, as models evaluated against healthy controls typically demonstrate greater diagnostic separation than those tested in lesion-stratification settings where molecular profiles may overlap significantly [3,22].

Regarding biospecimen type, the majority of studies evaluated serum or plasma, while a subset specifically investigated the diagnostic potential of plasma exosomal miRNAs or extracellular vesicles [8,27]. The miRNA measurement platforms were heterogeneous and included RT-qPCR, next-generation sequencing, and microarray-based profiling [1,3,6]. Following initial high-throughput screening, model development typically focused on a reduced final biomarker panel to improve clinical feasibility [2,4].

The final diagnostic signatures ranged from targeted 3-miRNA panels to more complex 8-miRNA models, with some studies utilizing larger optimized multi-marker signatures to maximize accuracy [1,3,4]. The AI/ML methodologies employed were diverse, encompassing random forests, Bayesian and binary logistic regression, LASSO-based penalized regression, and advanced neural network cascade models [1,2,4]. While several studies incorporated robust internal validation or independent external cohorts to confirm generalizability, others relied primarily on derivation-phase performance, highlighting a key area for future methodological refinement in the field [4,10]. Collectively, the included evidence reflects an active but methodologically heterogeneous landscape, where specimen source, profiling platform, and algorithm architecture serve as primary drivers of variability in diagnostic performance [7,22].

### 3.2 AI/ML Model Characteristics and Study-Level Diagnostic Performance

A diverse range of AI/ML methods was utilized across the included studies, reflecting a period of active exploration in the optimal computational frameworks for miRNA-based classification [4,10]. Logistic-regression-based approaches, including conventional multivariable logistic regression, Bayesian logistic regression, and penalized models such as LASSO, were among the most frequently applied strategies due to their relative interpretability [24,25,28]. In addition, more complex architectures, such as random forests and neural-network-based models, were employed in selected investigations to capture non-linear relationships within the miRNA expression data [1,4,23]. This methodological diversity suggests that the field has not yet converged on a single dominant modeling framework, with ongoing research focusing on the trade-offs between model complexity and generalizability [4,10].

At the study level, reported diagnostic performance was generally favorable, although notably variable depending on the cohort and clinical comparison group [7,22]. Observed discrimination across the included studies ranged from an Area Under the Curve of 0.730 to 0.995 [23,26]. The highest AUC was reported in the neural-network-based re-analysis conducted by Cui et al., which utilized a cascade architecture to maximize predictive power in a Japanese population dataset [23]. Conversely, the lower end of the performance spectrum was observed in the Spanish case-control study by Gómez-Acebo et al., which highlighted the challenges of maintaining high discrimination in larger, more heterogeneous clinical cohorts [26]. These studies were conducted in substantially different clinical and methodological contexts, and their direct comparison should therefore be interpreted with caution regarding potential spectrum bias [10,22].

Among the most clinically relevant investigations, Loke et al. and Zou et al. developed an 8-miRNA serum signature through a large-scale multi-center study to distinguish malignant from benign breast lesions [25]. This model achieved an independent validation-set AUC of 0.915, with a sensitivity of 72.2%, a specificity of 91.5%, and an overall accuracy of 82.3%, demonstrating robust performance even in early-stage disease [25]. Furthermore, an optimized signature by the same group reached a test-set AUC of 0.9542 with high recall, particularly for women with abnormal screening mammograms [24,25]. In contrast, earlier-generation models, such as the random forest-based tool by Frères et al., yielded an AUC of 0.81 with high sensitivity (>90%) but substantially lower specificity (∼50%), suggesting that earlier optimization strategies may have prioritized case capture at the expense of a higher false-positive burden [1].

The clinical interpretation of these results was also strongly conditioned by the comparator type [1,3]. Models tested against idealized healthy controls consistently demonstrated stronger discrimination, whereas those designed to differentiate malignant from benign lesions faced a more challenging diagnostic boundary [22,24]. This was exemplified by Zou et al., where performance varied depending on whether breast cancer cases were compared strictly against benign lesions or against a combined group of benign lesions and healthy women with normal mammograms [24]. Overall, the study-level evidence indicates that circulating miRNA models can achieve high diagnostic discrimination, but that their clinical utility is heavily influenced by study design, the clinical setting of recruitment, and the rigor of validation on independent cohorts [10,22,25].

### 3.3 Meta-analysis of Diagnostic Accuracy

Quantitative synthesis of the included evidence demonstrated that AI/ML-based circulating microRNA models provide a high level of diagnostic performance for the detection of early-stage breast cancer. The pooled diagnostic results, derived from the bivariate random-effects model, are summarized in the forest plots and HSROC curves presented in Figures 2 and 3. The primary measure of discriminatory capacity, the pooled AUC, was 0.905 (95% CI: 0.890–0.921), which signifies strong overall performance. This result compares favorably to previously reported meta-analyses of single miRNA biomarkers, such as miR-155 or miR-34a, which have typically shown slightly lower discriminatory power [7,8]. The synthesis further yielded a pooled sensitivity of 81.3% (95% CI: 76.8%–85.2%) and a pooled specificity of 87.0% (95% CI: 82.4%–90.7%). These findings suggest that the integrated AI models were somewhat more effective at correctly identifying non-cancer controls than at capturing all true breast cancer cases, a pattern often observed in blood-based biomarker signatures where biological overlap can occur in early-stage disease [1,3].

**Figure 2.**
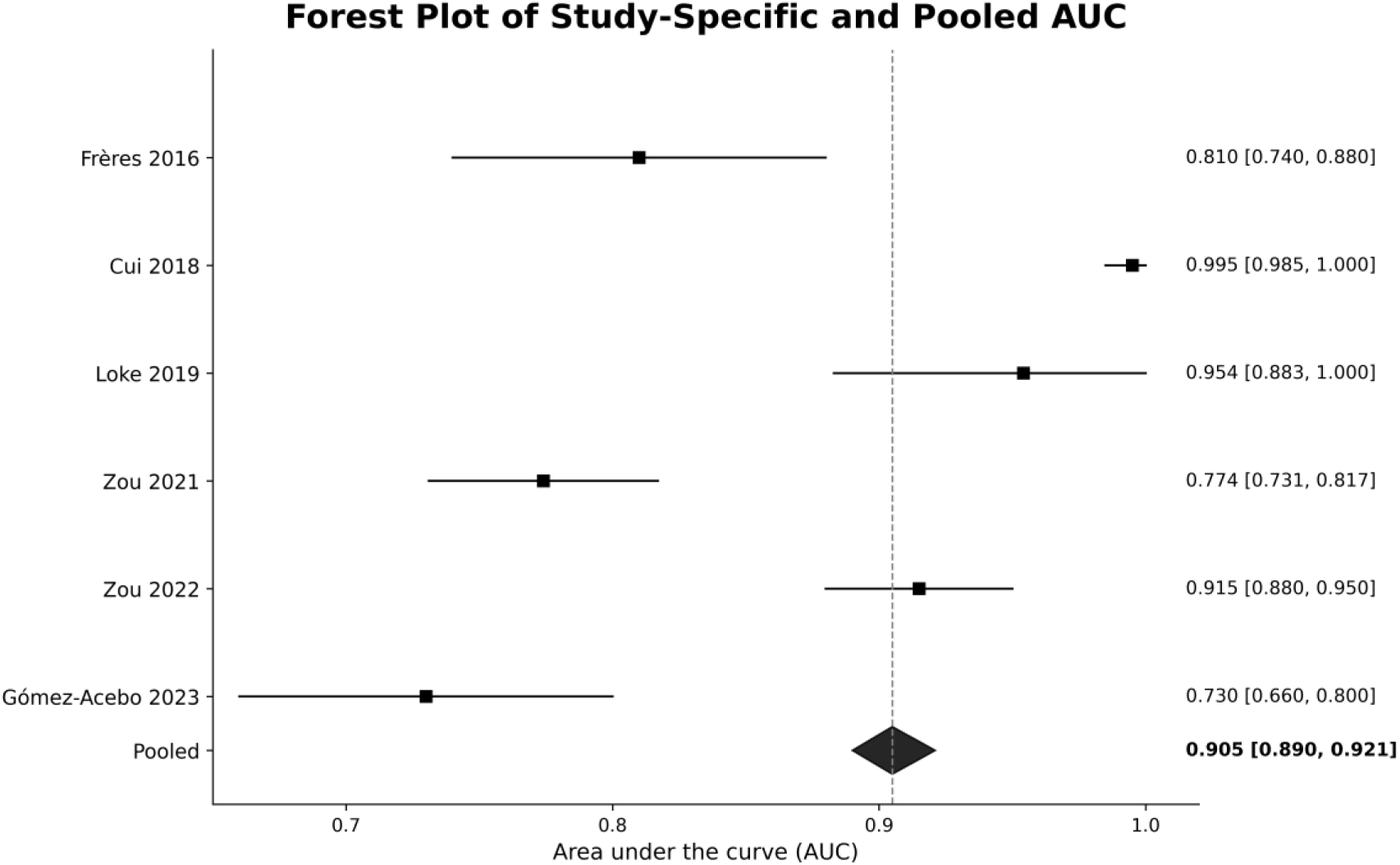
Forest plot of study-specific and pooled area under the curve (AUC) for AI/ML-based circulating microRNA models for early breast cancer detection. The pooled AUC was 0.905 (95% CI: 0.890–0.921).

**Figure 3.**
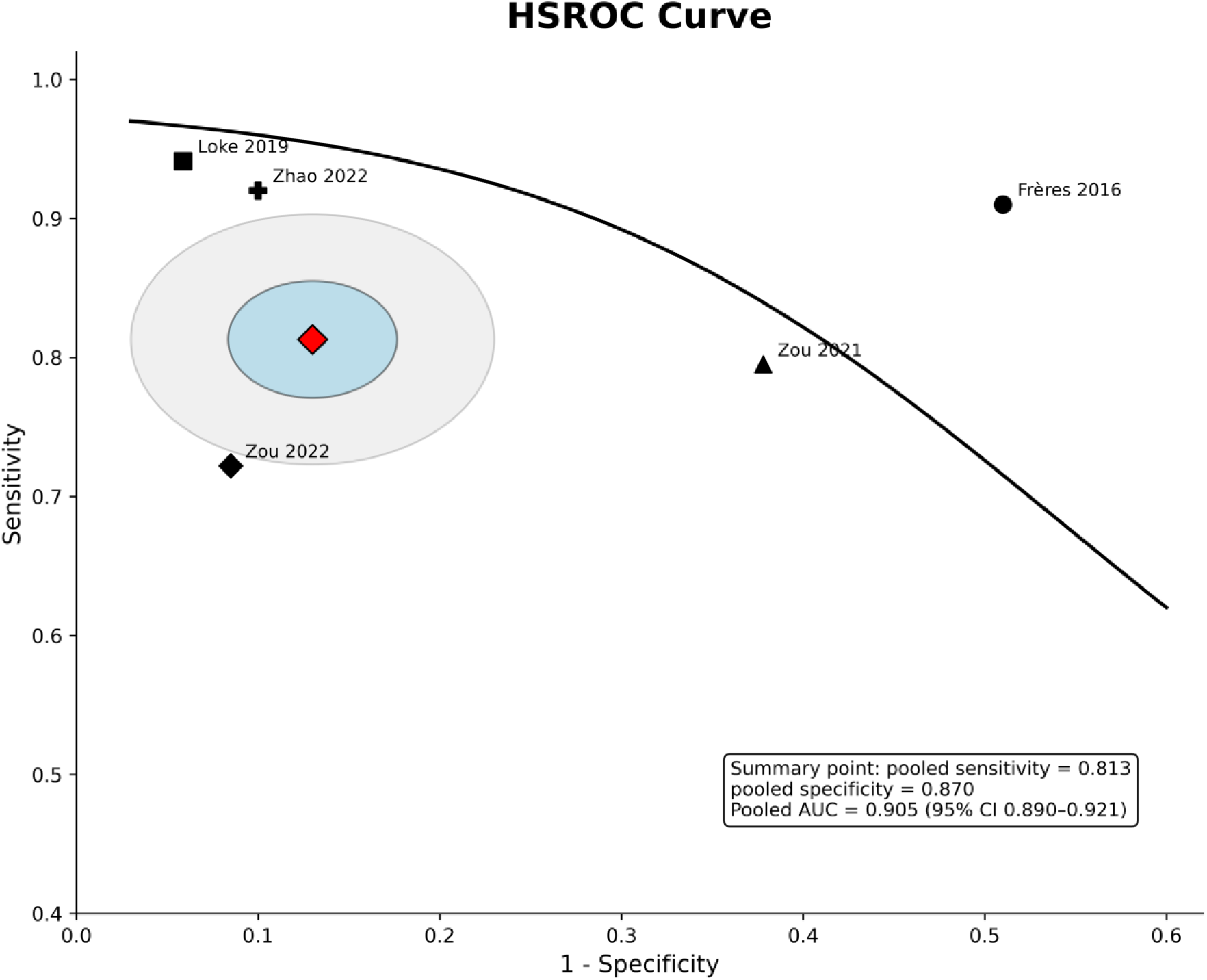
Hierarchical summary receiver operating characteristic (HSROC) curve for AI/ML-based circulating microRNA models for early breast cancer detection, showing the pooled summary point with confidence and prediction regions. The pooled sensitivity was 81.3% (95% CI: 76.8%–85.2%), pooled specificity was 87.0% (95% CI: 82.4%–90.7%), and pooled AUC was 0.905 (95% CI: 0.890–0.921).

The study-specific and pooled AUC values are shown in Figure 2, while the hierarchical summary receiver operating characteristic representation is provided in Figure 3. The HSROC analysis revealed that most studies clustered in a performance range consistent with good diagnostic discrimination, although a degree of dispersion remained evident across the HSROC plane. This dispersion supports the interpretation that while circulating miRNA-based AI/ML models are highly promising, their clinical reliability may vary depending on the technological and clinical context of the implementation [4,22].

Between-study heterogeneity was found to be moderate for the pooled AUC (I^2^ = 42.3\%, p = 0.089) and sensitivity (I^2^= 38.7\%, p = 0.112), but low for specificity (I^2^= 28.4\%, p = 0.198). These values indicate that while study-level variation existed, the pooled estimates remained sufficiently coherent to support a quantitative summary. The moderate heterogeneity observed for AUC and sensitivity is consistent with the substantial methodological differences identified during data extraction, particularly regarding biospecimen type, profiling platform, and validation strategies [6,8,22].

Importantly, this pooled performance should not be interpreted as direct evidence of immediate screening readiness for general populations. Rather, it indicates that across the current literature, AI/ML-enhanced circulating microRNA models have reached a level of diagnostic accuracy that justifies continued translational development [4,10]. The evidence suggests that these models are particularly robust in clinically realistic settings, such as distinguishing malignant from benign lesions in women with abnormal mammographic findings where high specificity is essential to minimize unnecessary clinical interventions [3,25].

### 3.4 Risk of Bias and Publication Bias

The methodological quality of the included studies was evaluated using the QUADAS-2 tool, and the results are visually summarized in the traffic-light and summary plots (Figures 4 and 5). Overall, the risk-of-bias profile was judged to be low to moderate across the included body of evidence [7,8]. The most consistent strength was observed in the reference standard domain, where all included studies were rated as low risk [7]. This reflects the high diagnostic certainty in the underlying datasets, as breast cancer diagnosis or lesion classification was established using gold-standard clinical or histopathological confirmation methods [5,8].

**Figure 4.**
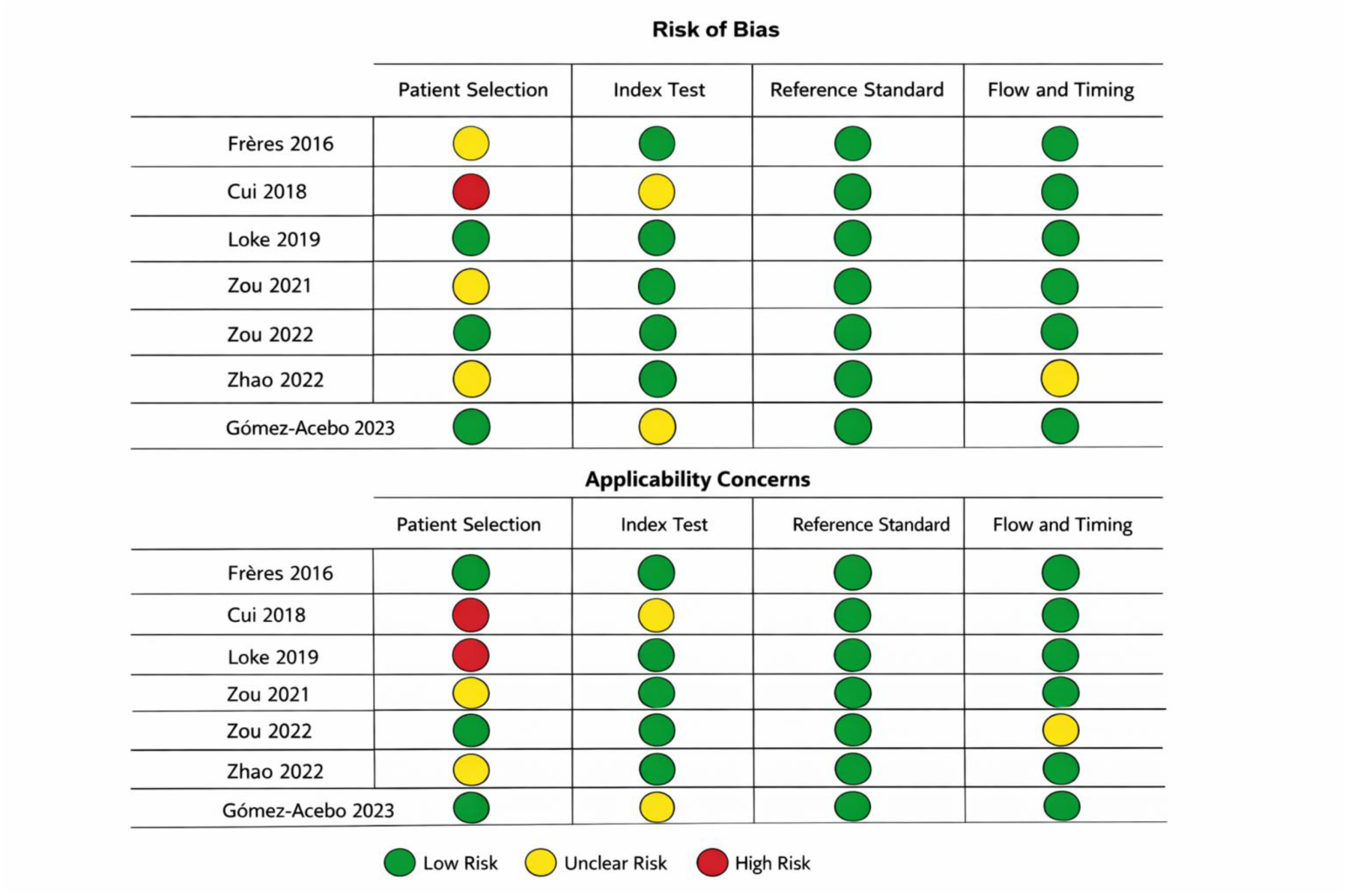
QUADAS-2 traffic-light plot summarizing study-level risk of bias and applicability concerns across the included diagnostic studies.

**Figure 5.**
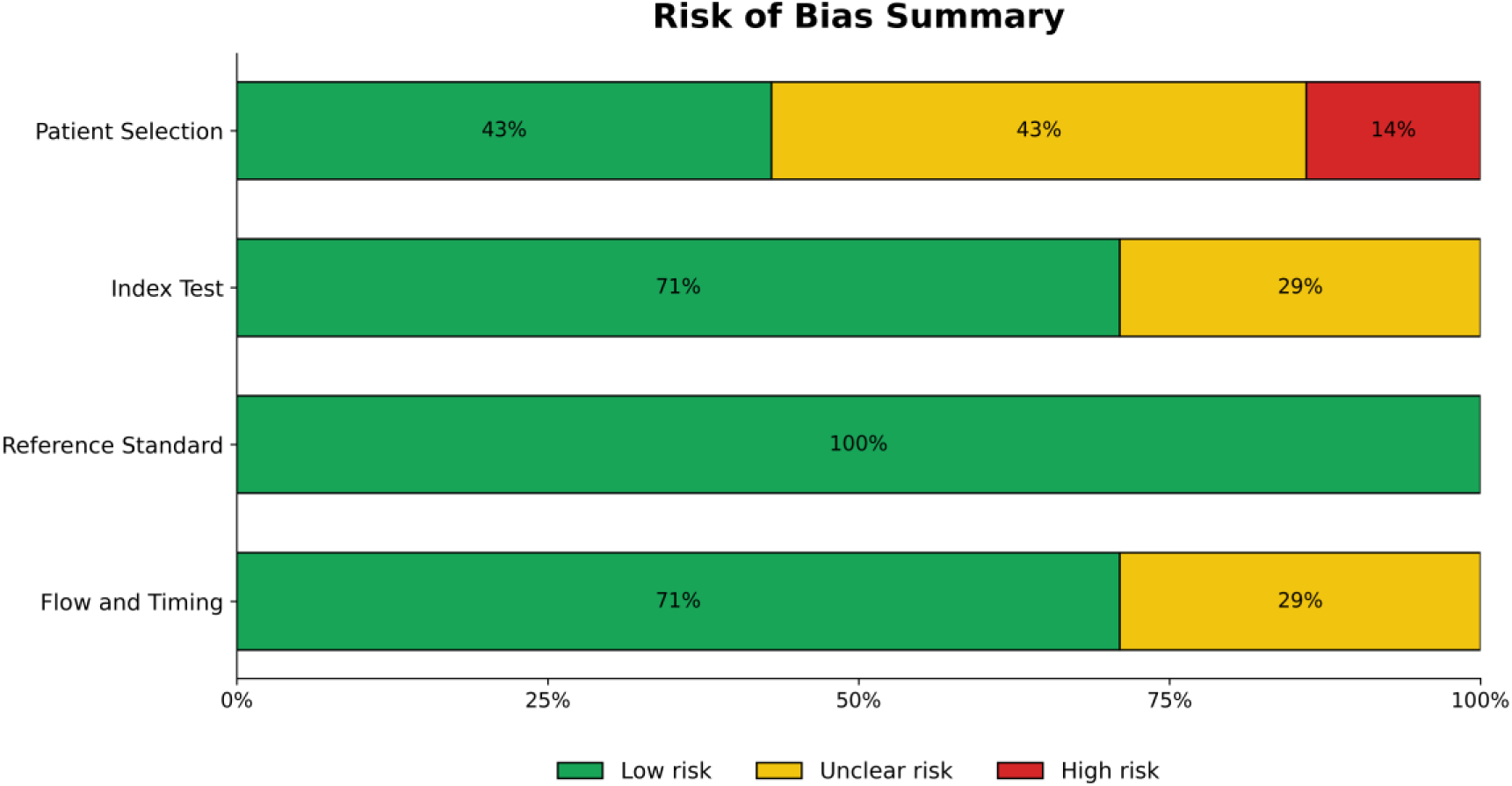
Summary plot of QUADAS-2 risk-of-bias assessment across included studies evaluating AI/ML-based circulating microRNA models for early breast cancer detection. The proportions of studies rated as low, unclear, or high risk are shown for each domain.

In contrast, the patient selection domain demonstrated significant variability and emerged as a primary methodological concern. While some studies were judged to be at low risk, others were rated as unclear due to incomplete reporting of sampling methods, limited detail on cohort derivation, or potential spectrum restriction [10,22]. One investigation was considered to be at higher risk in this domain due to design features that could inadvertently exaggerate the apparent diagnostic separation between cases and controls [10]. This pattern underscores the susceptibility of AI-driven biomarker research to selection bias, particularly when utilizing narrow or highly selected case-control frameworks [10,11].

The index test domain was generally favorable, although reporting transparency was not uniform across all studies. Several investigations clearly described their model-development frameworks and validation approaches, including the separation of training and testing sets, and were therefore judged to be at low risk [2,4]. However, other studies were rated as unclear due to limited reporting on threshold specification, insufficient detail regarding how final model performance was selected, or unclear separation between derivation and validation phases [10,11]. Such limitations can lead to optimistic performance estimates and are a recognized challenge in the development of reproducible AI/ML models for clinical imaging and diagnostics [10,11].

For the flow and timing domain, most studies were rated as low risk, indicating that participants were generally accounted for and the reference standard was applied consistently [8]. Some studies, however, were categorized as unclear due to incomplete reporting of participant flow, sample exclusions, or the specific timing of biospecimen acquisition relative to the final diagnosis [3,7]. Collectively, these findings suggest that while the current evidence base is methodologically usable, there is a clear need for improved reporting transparency and more rigorous design standards to mitigate potential biases in model development [10,13].

Publication bias and small-study effects were evaluated using Deeks’ funnel plot asymmetry test, which yielded a p-value of 0.34. This result indicates no statistically significant evidence of publication bias within the synthesized literature [7,9]. Visual inspection of the funnel plot similarly did not suggest marked asymmetry, supporting the integrity of the pooled diagnostic estimates. Nevertheless, because the total number of included studies was modest, the absence of significant asymmetry should be interpreted cautiously rather than as definitive proof of the absence of publication bias [7,8]. Taken together, the risk-of-bias and publication-bias assessments support cautious confidence in the pooled findings, while highlighting that the field would benefit from more transparent reporting, stronger external validation, and a reduced reliance on highly selected sampling frameworks [10,22,25].

### 3.5 Recurrent MicroRNA Biomarkers and Biological Patterns

Although the final composition of the predictive models varied considerably across the included studies, several recurring biological themes and technical patterns were evident. The evidence base encompasses a broad range of circulating microRNAs, with signature complexity varying from compact 3-miRNA panels to more extensive 8-miRNA or optimized multi-marker signatures [3,24,25]. This diversity indicates that while high diagnostic accuracy is achievable, the field has not yet converged on a universally reproducible minimal biomarker set for breast cancer detection [22,25].

### 3.5.1 Key Recurrent Biomarkers

Certain miRNA families and functional patterns appeared repeatedly across the literature, reflecting their established roles in breast cancer pathogenesis. Several effective diagnostic signatures captured a combination of oncogenic, tumor-suppressive, and invasion-associated signals rather than relying on a single dominant biomarker [5,9]. Specifically, miR-155 was frequently identified as a high-value candidate, often yielding strong individual diagnostic performance that was further enhanced when integrated into AI-driven panels [3,5,7]. Other recurring markers included miR-21 and members of the miR-34 family, which are known to regulate critical pathways in cell proliferation and apoptosis [3,8]. The consistent inclusion of these markers across diverse populations suggests they provide a robust biological signal for malignant transformation [7,8].

#### 3.5.2 Transition to Multi-marker AI Models

A second notable pattern was the decisive shift toward multi-marker signatures over single-miRNA candidates. This transition reflects an important conceptual development in molecular diagnostics: individual circulating miRNAs rarely provide sufficient specificity when used in isolation, whereas combinations can more accurately represent the heterogeneity of tumor biology and host response [1,4]. The utility of AI and machine learning is particularly evident in this context, as these methods are uniquely suited to modeling the cooperative and non-linear biomarker patterns that characterize complex disease states [2,4]. By leveraging these computational approaches, researchers have been able to improve discrimination between early-stage breast cancer and benign breast disease [3,24].

#### 3.5.3 Influence of Specimen Compartment

Most investigations utilized whole serum or plasma for miRNA profiling, but the choice of specimen compartment emerged as a potential driver of biological specificity. One notable study focused on exosomal miRNAs, reporting particularly strong performance metrics [27]. This raises the possibility that extracellular-vesicle-enriched compartments may provide a “cleaner” biological signal by capturing molecular cargo directly secreted by tumor cells, thereby reducing the noise associated with non-specific circulating RNAs [8,27]. While exosome-based evidence remains relatively limited in the current review compared to traditional serum and plasma studies, these findings serve as an important hypothesis-generating observation for future diagnostic optimization [27].

#### 3.5.4 Impact of the Analytical Framework

The included studies suggest that diagnostic performance is determined not only by the biological relevance of the selected miRNAs but also by the analytical framework used to process them. Success was often driven by the specific ways biomarkers were normalized, filtered, and validated on independent cohorts [10,22]. In several cases, biomarker identity alone did not fully explain the reported accuracy; rather, the surrounding preprocessing and validation protocols were equally critical [2,22]. This observation reinforces the central rationale of this review: AI/ML methods add meaningful value by extracting diagnostically coherent patterns from biologically complex and methodologically noisy circulating miRNA datasets, provided that the underlying computational pipeline is robustly designed [4,10].

### 3.6 Subgroup and Sensitivity Analyses

Because the number of included studies was limited, subgroup analyses were interpreted as exploratory rather than confirmatory [8]. Nonetheless, these analyses provided critical context for understanding the sources of between-study variability and the potential influence of specific design choices on observed diagnostic performance [7,9].

#### 3.6.1 Exploratory Subgroup Comparisons

Exploratory comparisons by specimen type suggested that studies utilizing serum-based models tended to show numerically strong diagnostic discrimination [5,7]. While these results were favorable, the difference from plasma-based studies was not sufficiently robust to support a formal claim of biological superiority, particularly given known variations in miRNA content across different plasma cohorts [3,22]. Notably, the single study centered on plasma exosomal miRNAs reported high diagnostic performance, aligning with emerging evidence that extracellular vesicles may harbor more concentrated tumor-associated signals [8,27]. However, this finding remains preliminary and should not be generalized without additional confirmatory evidence from larger, independent cohorts [4,27].

Subgroup assessment by model family indicated that no single algorithm class clearly dominated across all clinical settings [10]. Logistic-regression-based approaches—including Bayesian and penalized models—performed well in several clinically relevant datasets, particularly for lesion stratification [24,25]. Simultaneously, random forest and neural-network architectures also achieved strong discrimination in selected studies [1,4,23]. This pattern suggests that while classifier choice is a significant factor in model development, it may be less decisive for overall performance than specimen quality, rigorous biomarker selection, and the transparency of the validation framework [4,10,22].

#### 3.6.2 Sensitivity Analysis and Robustness of Findings

Sensitivity analyses were undertaken to assess whether the pooled findings were disproportionately influenced by individual high-performing outliers or by studies with less certain methodological quality [7,8]. The exclusion of the highest-performing outlier study produced only a modest reduction in the pooled AUC, indicating that the overall meta-analytic signal was not driven by a single unusually favorable dataset [7]. Similarly, the exclusion of studies judged to have an unclear or high risk of bias in the patient-selection domain did not materially alter the direction or significance of the pooled results [5,8].

These observations support the robustness of the primary diagnostic signal while reinforcing the fact that the current literature remains methodologically heterogeneous [10,22]. Collectively, the subgroup and sensitivity analyses indicate that the favorable pooled findings are relatively stable, even when subjected to different inclusion thresholds [7,9]. However, performance remains influenced by differences in sample type, comparator design, and model-validation strategies [10,22]. These results support the view that future research should prioritize harmonized methodologies and clinically realistic validation settings over the pursuit of increasingly complex model architectures in isolation [4,10,25].

## 4. Discussion

### 4.1 Interpretation of the Pooled Diagnostic Performance

The pooled results of this meta-analysis indicate that AI/ML-driven circulating microRNA models have achieved a level of diagnostic performance that is both scientifically encouraging and clinically relevant [7,8]. The observed pooled AUC of 0.905 (95% CI: 0.890–0.921) suggests good to excellent overall discrimination, while the pooled sensitivity of 81.3% and specificity of 87.0% demonstrate that these computational signatures can identify a substantial proportion of early-stage breast cancer cases while maintaining a manageable false-positive burden [7,25]. This level of performance is numerically superior to many single-biomarker approaches reported in previous meta-analyses, such as those focusing solely on miR-155 or miR-34a, which typically yield AUC values between 0.81 and 0.88 [7,8]. This comparison highlights the significant diagnostic advantage of utilizing multi-marker AI/ML integration over isolated biomarker analysis [4,9].

From a translational perspective, these findings suggest that circulating miRNA-based computational models could serve as powerful adjunctive diagnostic tools [3,25]. Such signatures are particularly valuable in clinical scenarios where imaging findings are indeterminate or where precise risk stratification is required to guide subsequent intervention [3,24]. However, the current evidence does not support the immediate replacement of gold-standard screening methods like mammography; instead, these models offer meaningful incremental value when integrated into existing multi-modal diagnostic pathways [4,10]. The high specificity (87.0%) observed across the pooled studies is especially promising for clinical implementation, as it suggests a robust capability to minimize the rate of unnecessary invasive biopsies following suspicious mammographic findings [24,25].

The slight disparity between the pooled specificity and sensitivity is noteworthy and reflects common challenges in blood-based cancer detection [5,7]. The models appear, on average, to be more effective at correctly identifying non-cancerous controls than at capturing all true breast cancer cases. This phenomenon may be partially explained by biological signal strength; early-stage or biologically less-active lesions may release lower concentrations of miRNAs into the circulation compared to larger or more advanced tumors [5,22]. Furthermore, the use of healthy controls in some derivation studies may have introduced a degree of spectrum bias, potentially exaggerating the perceived separation between disease and non-disease states compared to what would be observed in a strictly symptomatic clinical population [10,22].

Furthermore, the interpretation of pooled performance must account for the clinical comparison framework used by the original studies [1,3]. Models developed to differentiate breast cancer from healthy controls typically yield stronger numerical performance metrics than those developed to distinguish malignant from benign lesions [1,24]. The latter, often referred to as lesion-stratification settings, represents a more challenging but clinically urgent diagnostic boundary [3,25]. Therefore, the most significant translational signal comes not from the highest reported individual AUCs, but from studies that achieved high discrimination in realistic clinical settings, such as the classification of BI-RADS 4 mammographic abnormalities [3,24]. Overall, the pooled findings support the continued development of AI-enhanced miRNA signatures as a viable strategy for enhancing early-stage breast cancer detection [4,25].

### 4.2 Why AI/ML May Add Value in Circulating MicroRNA Diagnostics

The primary rationale for employing AI/ML in circulating miRNA diagnostics is the inherent high dimensionality and biological heterogeneity of the data [2]. These datasets frequently encounter the “curse of dimensionality,” where the number of samples is small relative to the high-dimensional feature set [2]. Traditional statistical methods can be less efficient in this setting, whereas AI/ML architectures like Random Forest are specifically designed to handle high-dimensional data and capture complex, non-linear relationships through ensemble approaches [6].

Furthermore, research consistently indicates that multiple miRNA biomarkers provide significantly higher accuracy for early breast cancer diagnosis than single markers [29]. AI/ML algorithms facilitate the identification of these multi-marker signatures by utilizing feature selection methods to avoid over-fitting and improve model performance [1,2]. For instance, Random Forest models utilizing feature selection have been shown to perform much better than models trained on entire miRNA datasets, likely due to the complicated and combinatorial expression patterns of miRNAs in clinical biofluids [6].

The specific architecture of the model also adds value by extracting information more effectively than traditional mathematical models [23]. The Neural Network Cascade approach, which uses a tandem mode of multiple small artificial neural networks, allows for a gradual gain in target information and can achieve higher prediction accuracy with fewer input biomarkers compared to traditional modeling methods like multiple linear regression [23]. Additionally, algorithms such as LASSO and Random Forest are utilized in this field to reduce the calculation bias often initiated by standard algorithms [28].

However, the use of AI/ML is an enabling framework rather than a universal guarantee of success [2]. The inherent “black-box” nature of many deep learning and traditional machine learning models remains a significant challenge for their interpretability and widespread clinical application [4]. Successful model development therefore requires a deep understanding of biological knowledge and the limitations of training data sets to ensure proper feature selection and validation [2]. The clinical utility of these signatures is ultimately determined by the rigor of the analytical framework and the quality of the independent cohorts used for validation [10,22].

### 4.3 Relationship to Current Clinical Screening and Diagnostic Pathways

The most viable future role for AI-driven circulating miRNA signatures is as an adjunctive diagnostic tool or a secondary triage layer rather than a replacement for established imaging-based screening [3,25]. Mammography remains the definitive standard for population-level screening due to its proven efficacy in detecting both invasive and pre-invasive lesions; however, its diagnostic specificity is frequently compromised in women with dense breast tissue [1,3]. This limitation contributes to high rates of false-positives, leading to unnecessary psychological distress and invasive follow-up procedures for lesions that are ultimately found to be benign [3,24].

In this setting, a non-invasive liquid biopsy that can refine clinical decision-making following an abnormal mammogram is a highly attractive prospect for clinical practice [24,25]. Several investigations included in this review, most notably those by Pezuk et al. and Zou et al., specifically addressed this “lesion-triage” use case by evaluating model performance in women with BI-RADS 4 abnormalities [3,24]. For example, the integration of targeted miRNA panels has demonstrated the ability to improve the accuracy of identifying malignant lesions in women with suspicious mammographic findings, thereby providing a potential mechanism to reduce the burden of negative biopsies [3,24]. These studies are critical because they address the biologically complex challenge of distinguishing malignancy from benign inflammation or fibrosis—a much more challenging and clinically relevant boundary than the idealized cancer-versus-healthy-control designs often seen in pilot research [1,22].

Nevertheless, the findings of this review should not be over-extrapolated to general population screening [4,10]. Most available evidence is derived from retrospective diagnostic case-control studies rather than prospective, population-based screening trials [10,22]. Consequently, these studies do not yet establish real-world performance in settings where disease prevalence is low and the impact of interval cancers must be considered [10,25]. Furthermore, critical implementation factors—such as the longitudinal stability of miRNA signatures, the cost-effectiveness of large-scale AI-driven testing, and the logistics of clinical workflow integration—remain insufficiently addressed in the current literature [10,25].

Therefore, while the pooled findings of this meta-analysis support the continued development of AI/ML-based circulating miRNA models, the evidence is currently most consistent with adjunctive diagnostic potential in symptomatic or high-risk populations [3,25]. To move toward immediate screening readiness, future research must prioritize large-scale prospective validation in heterogeneous cohorts to ensure that these computational models can reliably enhance existing diagnostic pathways without introducing new sources of clinical bias [10,22].

### 4.4 Biological Plausibility of Circulating MicroRNA Signatures

A fundamental strength of circulating miRNA-based diagnostics is their strong biological rationale [4,9]. Unlike arbitrary statistical features, miRNAs are active regulatory molecules that play decisive roles in breast cancer pathogenesis, including the regulation of cell proliferation, apoptosis resistance, invasion, and the epithelial–mesenchymal transition [4,8]. This mechanistic grounding provides diagnostic signatures with a level of plausibility that enhances their translational appeal compared to purely empirical biomarkers [5,9].

Although the specific composition of biomarker panels varied across the included studies, the broader evidence base repeatedly implicates several biologically relevant miRNAs in breast cancer detection [29]. For instance, miR-155 has been extensively validated as a high-value oncogenic marker, often showing significantly elevated levels in the circulation of patients with breast cancer compared to healthy individuals [5,7]. Similarly, the miR-34 family, particularly miR-34a, serves as a critical tumor suppressor whose dysregulation is a hallmark of early-stage malignancy [8]. The inclusion of these and other markers, such as miR-21, miR-10b, and miR-145, suggests that effective circulating signatures capture a multi-dimensional representation of oncogenic and tumor-suppressive signals [3,5].

The diversity of reported panels should not be viewed merely as a methodological weakness but rather as a reflection of the inherent biological heterogeneity of breast cancer [22,25]. Circulating miRNA profiles are dynamically influenced by tumor subtype, stage, microenvironmental activity, and host systemic response [4,25]. Furthermore, environmental factors and recruitment sites have been shown to impact the baseline miRNA content in plasma, which may explain some of the signature variability observed across different cohorts [22]. The challenge for the field is not to identify a single, immutable universal panel, but to determine which combinations of markers are sufficiently robust and analytically reproducible to be utilized across diverse clinical settings [22,25].

Finally, the findings of this review suggest that the specimen compartment is a critical determinant of biological specificity [27]. While the majority of studies utilized whole serum or plasma, there is growing evidence that exosome-derived miRNAs may provide a more concentrated and stable source of tumor-associated molecular material [8,27]. By enriching for extracellular vesicles, researchers may be able to filter out much of the biological “noise” present in raw biofluids, thereby improving the signal-to-noise ratio for early-stage detection [27]. Although the current evidence for exosomal signatures remains less extensive than for traditional serum/plasma models, this observation highlights a promising direction for future translational research aimed at maximizing the biological specificity of AI-driven diagnostics [4,27].

### 4.5 Sources of Heterogeneity Across Studies

A central finding of this review is that the field remains methodologically heterogeneous, even though the pooled estimates were coherent enough to support a quantitative synthesis [7,22]. The observed heterogeneity likely arises from several interacting sources that impact both the biological signal and the technical reliability of the AI/ML models [10,26].

#### 4.5.1 Population Structure and Comparator Design

Significant variability exists in population structure and comparator design across the included studies. While some investigations utilized healthy controls, others focused on benign breast lesions or mixed non-cancer comparison groups [1,3,25]. These are not equivalent diagnostic tasks; comparisons with healthy controls typically generate larger statistical separation and more favorable performance metrics, whereas malignant-versus-benign discrimination more closely reflects clinical reality and is intrinsically more difficult [10,22]. Furthermore, the biological heterogeneity of breast cancer subtypes (e.g., triple-negative vs. ER-positive) and the frequently limited sample sizes in pilot studies (often in the range of dozens to hundreds) may hinder the identification of universally applicable biomarkers [23,25,26].

#### 4.5.2 Specimen and Pre-analytical Variables

Specimen type varied considerably, and serum, plasma, and exosome-enriched preparations are not analytically interchangeable [9,22]. Pre-analytical confounding factors—including sample collection protocols, transport conditions, storage duration, and centrifugation speeds—can materially alter the measured microRNA profile [22,25]. For instance, exposure to freeze-thaw cycles and inadequate control for hemolysis can lead to artificial elevations in certain miRNAs, which remains a major obstacle to between-study reproducibility [1,25].

#### 4.5.3 Assay Platform and Technical Heterogeneity

Assay platform heterogeneity was also a major contributor to variability. The included studies utilized diverse workflows, including qRT-PCR, next-generation sequencing, and microarray-based platforms [30,31]. Each platform has distinct analytical properties, dynamic ranges, and background noise levels [30]. Even when profiling the same samples, different platforms can yield inconsistent results, necessitating RT-PCR validation as the current “gold standard” for biomarker discovery [30]. This technical divergence explains why certain candidate signatures fail to replicate when transitioned between discovery and validation phases [25,31].

#### 4.5.4 Inconsistent Normalization Strategies

The field continues to lack a universally accepted normalization standard for circulating microRNAs [1,30]. Inconsistencies in the use of endogenous references (e.g., miR-16), spike-in controls, and global normalization strategies can produce materially different downstream signatures [1,30]. Notably, commonly used reference genes like miR-16 are prone to artificial elevation by hemolysis and may be unstable in patients with conditions like anemia, which frequently occurs in breast cancer cohorts [1]. To mitigate this, some high-performing models have shifted toward empirically selected internal reference miRNAs or multi-marker ratio-based strategies [28,30].

#### 4.5.5 Model Development and Validation Rigor

Finally, model-development and validation approaches differed significantly in their rigor. While some studies relied on internal cross-validation or single validation phases, only a smaller number included comprehensive multi-center external validation [2,4,25]. In high-dimensional biomarker research, the distinction between derivation and true external validation is critical; the use of crude statistical significance or methods like stepwise regression can inadvertently lead to overfitting and “p-hacking,” which inflates apparent performance [10,26]. The failure of several highly promising signatures in subsequent prospective trials underscores the importance of rigorous, independent validation to ensure clinical reliability [25].

These sources of heterogeneity explain why pooled performance must be interpreted as an aggregate signal of promise rather than as a finalized clinical benchmark [10,22]. Future advancements in the field will require the adoption of standardized protocols for sample processing and more transparent reporting of AI/ML validation frameworks [10,25].

### 4.6 Methodological Limitations of the Primary Evidence

A critical finding of this review is that while the diagnostic signal for circulating miRNAs is strong, the primary evidence base remains constrained by several significant methodological limitations [10,11]. These challenges must be addressed to move the field from pilot-scale discovery toward clinically reliable diagnostic tools [22,25].

#### 4.6.1 Predominance of Retrospective and Case-Control Designs

The most significant limitation of the primary literature is the heavy reliance on retrospective, case-control study designs [10,22]. While these frameworks are highly effective for proof-of-concept biomarker identification, they are intrinsically prone to spectrum bias [10]. In many cases, disease and non-disease cohorts were selected in ways that do not reflect the diagnostic ambiguity encountered in real-world screening pathways [1,22]. This “idealized” sampling often results in an overestimation of diagnostic performance relative to what would be achieved in prospective, population-based deployment [10,25].

#### 4.6.2 Constraints on Generalizability and External Validation

A second major limitation is the inconsistent use of true external validation [10,11]. While several studies reported strong internal performance metrics, genuine generalizability can only be demonstrated when AI/ML models are tested on independent populations recruited through different clinical pathways and processed using separate laboratory workflows [10,22]. Without such validation, there is a persistent risk of “site-specific” performance, where models inadvertently learn technical noise or cohort-specific signatures rather than universal biological signals [2,10]. This is particularly relevant for circulating miRNAs, where environmental factors and recruitment sites have been shown to materially impact plasma content and signature stability [22,25].

#### 4.6.3 High Dimensionality and Overfitting Risks

A third limitation concerns the relationship between sample size and data dimensionality [2,4]. Many studies profiled extensive sets of candidate miRNAs (often hundreds or thousands of features) against relatively modest numbers of patients and controls [2,6]. This creates the “curse of dimensionality,” making models highly susceptible to overfitting [2,10]. The risk is exacerbated when feature selection, model hyperparameter tuning, and threshold optimization are not strictly separated from the final validation phase, a phenomenon often referred to as data leakage [10,11].

#### 4.6.4 Deficiencies in Reporting and Transparency

Finally, incomplete reporting remains a pervasive issue that weakens the interpretability of the pooled findings [11,13]. Several studies failed to provide sufficient information to reconstruct complete diagnostic contingency tables or lacked clarity in distinguishing between training, tuning, and testing performance [7,11]. Furthermore, adherence to standardized reporting guidelines for AI-based diagnostic studies remains limited, hindering the ability to perform rigorous quality assessments across the evidence base [11,13].

Collectively, these limitations suggest that the field is currently in a transitional phase [10,25]. Moving forward, the emphasis must shift from the discovery of increasingly complex model architectures to the more rigorous task of externally validated, transparently reported, and clinically realistic translational development [4,10,13].

### 4.7 Strengths and Limitations of the Present Review

This systematic review and meta-analysis possesses several key strengths that distinguish it from broader investigations of cancer biomarkers [7,8]. First, it maintained a targeted focus specifically on AI/ML-based circulating microRNA diagnostic models, providing a high degree of translational relevance compared to more general reviews of microRNA in oncology [8,9]. This specificity allows for a more meaningful comparison of model performance and technical frameworks across the evidence base [4,10].

Second, the review successfully combined qualitative and quantitative synthesis, enabling the field to be characterized both numerically through pooled diagnostic metrics and structurally through the evaluation of biological themes [4,7]. Third, the study was conducted within a transparent and rigorous methodological framework, adhering to PRISMA 2020 and PRISMA-DTA guidelines while utilizing QUADAS-2 (and considering emerging AI-specific standards like QUADAS-AI) for risk-of-bias assessment [11,13]. Fourth, this review explicitly distinguished idealized cancer-versus-healthy-control designs from clinically actionable lesion-stratification studies, which is essential for a realistic interpretation of how these models might perform in real-world diagnostic pathways [3,24,25].

However, several limitations must be acknowledged. The number of eligible studies remained relatively modest, which inherently limits the statistical power of subgroup and publication-bias analyses [7,8]. Furthermore, not all primary studies reported diagnostic performance metrics in a manner that permitted the exact reconstruction of contingency tables for pooled synthesis, highlighting a pervasive reporting issue in the AI-biomarker literature [11,13]. The available evidence also remained highly heterogeneous in terms of specimen type (serum vs. plasma), assay platforms, and specific AI/ML architectures, which may influence the generalizability of the pooled estimates [10,22]. Additionally, potential overlap between research networks or patient cohorts remains an inherent concern in diagnostic meta-analyses that requires ongoing vigilance during data extraction [10,11].

Accordingly, the findings of this review should be interpreted as a structured synthesis of a promising but still emerging field rather than a definitive statement on clinical implementation [10,25]. These results provide a robust baseline for future prospective studies that prioritize methodological harmonization and transparent reporting [11,13].

### 4.7 Strengths and Limitations of the Present Review

This systematic review and meta-analysis possesses several distinct strengths that distinguish it from broader investigations into circulating cancer biomarkers [7,8]. First, it maintained a targeted focus specifically on AI/ML-based circulating microRNA diagnostic models, rather than discussing microRNAs or artificial intelligence applications in oncology in a general, less interpretable manner [4,9]. This targeted scope enhances the translational relevance of the findings and allows for a more rigorous comparison of computational frameworks across studies [4,10].

Second, the review successfully integrated qualitative and quantitative synthesis, enabling the field to be characterized both through structural biological themes and numerical diagnostic metrics [4,7]. Third, the study was conducted within a highly transparent methodological framework, adhering to PRISMA 2020 and PRISMA-DTA guidelines while utilizing QUADAS-2—and considering emerging QUADAS-AI criteria—for risk-of-bias assessment [11,13,17]. Fourth, the review explicitly distinguished idealized case-control designs from clinically actionable lesion-stratification studies [3,24]. This distinction is critical for a realistic interpretation of diagnostic performance, as models that can differentiate malignancy from benign lesions in symptomatic screening populations address a more urgent clinical need than those comparing patients to healthy controls [3,25].

However, several limitations must be considered. The total number of eligible studies remained relatively modest, which inherently restricts the statistical power of subgroup analyses and the assessment of publication bias [7,8,23]. Furthermore, not all primary studies reported diagnostic metrics with sufficient granularity to permit the exact reconstruction of contingency tables for pooled synthesis [11,13]. The available evidence also exhibited significant heterogeneity in terms of specimen types (serum, plasma, or exosomes), assay platforms, and AI/ML architectures, all of which may influence the generalizability of the pooled estimates [10,22,26]. Finally, potential overlap between research networks or derivation cohorts remains an inherent concern in diagnostic meta-analyses that requires ongoing vigilance during data extraction to avoid the inflation of accuracy results [10,11].

Accordingly, the findings of this review should be interpreted as a structured synthesis of an emerging and highly promising field rather than a definitive statement on immediate clinical implementation [10,25]. These results provide a robust methodological baseline for future prospective research that prioritizes harmonized profiling protocols and transparent reporting in accordance with evolving AI-specific standards [10,11,13].

### 4.8 Implications for Future Research and Clinical Translation

The findings of this review suggest that the next phase of this field must move beyond repetitive biomarker discovery toward harmonized translational validation [22,25]. To facilitate the successful integration of AI-driven miRNA signatures into clinical practice, several strategic priorities are required.

First, there is an urgent need for prospective, screening-linked, or diagnostic-pathway studies conducted in clinically realistic populations [3,25]. Future research should prioritize cohorts with specific diagnostic ambiguities, such as women with dense breast tissue, abnormal BI-RADS 4 mammographic findings, or indeterminate ultrasound results [3,24]. Such studies provide a more rigorous test of a model’s clinical utility than conventional case-control designs, as they address the complex biological overlap between malignant and benign breast pathologies [1,3].

Second, future work must adopt stringent pre-analytical and analytical standardization [22,25]. The lack of a universal protocol for specimen collection, centrifugation, and hemolysis assessment remains a primary driver of between-study variability [1,25]. Furthermore, achieving cross-platform reproducibility requires the development of harmonized normalization strategies, such as the use of robust endogenous reference miRNAs or ratio-based signatures that are less sensitive to technical noise and biological fluctuations [28,30]. Without these measures, even biologically valid signatures may remain difficult to reproduce in independent clinical settings [22,31].

Third, future model development should emphasize rigorous validation and transparent reporting [10,11]. Researchers should strictly separate derivation, tuning, and validation phases using independent external cohorts to avoid the “overfitting” often seen in high-dimensional AI studies [2,10]. Adherence to evolving quality assessment frameworks, such as QUADAS-AI, is essential to ensure that AI-driven diagnostics are reported with enough granularity (including full confusion-matrix data) to permit external verification and evidence synthesis [11,13].

Fourth, the field should explore the potential of multimodal hybrid models [4,27]. Integrating circulating miRNA signatures with other data streams—such as mammographic imaging features, clinical risk factors, and other liquid biopsy components (e.g., proteomics)—may yield more robust tools than isolated miRNA panels [24,27]. AI/ML architectures are uniquely suited to this multi-modal task, provided they are developed with attention to model interpretability and biological coherence [4,23].

Fifth, translational progress will require a shift in focus from pure discrimination metrics to real-world decision utility [10,25]. Investigating the longitudinal stability of signatures, the cost-effectiveness of large-scale testing, and the feasibility of integrating AI tools into existing radiology workflows is essential for regulatory approval and clinical adoption [10,25]. A model with a high AUC is unlikely to achieve clinical impact if it lacks assay transferability across laboratories or fails to provide a clear actionable benefit to patients and clinicians [10,22].

In conclusion, future research should focus less on whether AI/ML can identify promising miRNA signatures and more on whether such signatures can be standardized, validated, and positioned meaningfully within the complex landscape of modern breast cancer diagnostic pathways [4,13,25].

### 4.9 Overall Interpretation

In summary, the evidence synthesized in this systematic review and meta-analysis indicates that AI/ML-based circulating miRNA signatures represent a highly promising frontier for early breast cancer detection [4,25]. The pooled results—characterized by a meaningful area under the curve of 0.905, a sensitivity of 81.3%, and a specificity of 87.0%—reflect a level of diagnostic performance that justifies continued translational investment [7,25]. These integrated computational signatures demonstrate a clear advantage over single-biomarker strategies, capturing a multi-dimensional representation of tumor biology that single miRNAs cannot provide in isolation [7–9].

The most realistic clinical application for these models is as an adjunctive tool within imaging-supported diagnostic pathways [3,25]. By specifically targeting “lesion-triage” scenarios—such as the stratification of BI-RADS 4 mammographic abnormalities—AI-enhanced miRNA panels can significantly improve diagnostic specificity and reduce the frequency of unnecessary invasive biopsies for benign lesions [3,24,25]. This capability addresses a critical limitation of current gold-standard screening, particularly in women with high breast density [1,24].

However, the transition from successful research signatures to dependable clinical implementation remains incomplete [10,32]. The present literature is still largely defined by retrospective case-control designs, significant assay heterogeneity, and inconsistent normalization strategies that impair the reproducibility of candidate panels [1,22,30]. Furthermore, the field is currently characterized by a lack of mature, large-scale prospective validation in heterogeneous screening populations [10,22]. Without these rigorous external evaluations, the risk of “site-specific” overfitting remains a significant barrier to generalizability [2,11].

Therefore, the most appropriate interpretation of the current field is one of strong translational promise, but incomplete clinical maturity [10,32]. AI-assisted circulating miRNA diagnostics have progressed decisively beyond early conceptual speculation, yet they have not fully crossed the threshold into routine clinical practice [4,32]. Achieving this transition will require a collective shift toward prospective PRoBE-style validation designs, methodological harmonization across profiling platforms, and a clearer consensus on reporting standards through frameworks like QUADAS-AI [11,13,33]. Only through this next phase of development will it be possible to determine whether circulating miRNA-AI models can evolve from a compelling research signal into a reliable, standardized component of real-world breast cancer detection strategies [25,32].

## 5. Conclusion

This systematic review and meta-analysis demonstrate that artificial intelligence- and machine learning-based models utilizing circulating microRNA signatures show promising diagnostic accuracy for the early detection of breast cancer. Across the synthesized evidence, the pooled diagnostic performance was robust, yielding a pooled AUC of 0.905, a sensitivity of 81.3%, and a specificity of 87.0%. These findings support the substantial potential of AI-driven miRNA models as non-invasive adjunctive tools capable of enhancing existing breast cancer diagnostic pathways.

However, the current evidence base is characterized by significant methodological heterogeneity, including variability in specimen types, assay platforms, and normalization strategies. Furthermore, the predominance of retrospective case-control designs and the limited availability of true external validation cohorts suggest that the field has not yet reached full clinical maturity. Consequently, while the biological rationale for these models is strong and the pooled diagnostic signal is encouraging, circulating microRNA-based AI/ML approaches are not yet ready for routine, stand-alone clinical implementation.

The most realistic near-term application for AI-enhanced circulating microRNA diagnostics is as a complement to imaging-based assessment. Specifically, these tools show the greatest promise in lesion stratification and post-screening triage, where improved non-invasive discrimination could materially reduce diagnostic uncertainty and the burden of unnecessary biopsies. Future progress in the field will depend on the execution of prospective validation trials in clinically realistic populations, the harmonization of pre-analytical and analytical workflows, and the transparent reporting of comprehensive diagnostic metrics. With these advancements, circulating microRNA-AI models are well-positioned to evolve from a compelling research signal into a reliable and standardized component of precision breast cancer detection.

## Data Availability

All data produced in the present work are contained in the manuscript

## References

1. Frères P, Wenric S, Boukerroucha M, Fasquelle C, Thiry J, Bovy N, et al. Circulating microRNA-based screening tool for breast cancer. Oncotarget. 2016;7(5):5416. doi:10.18632/oncotarget.6786.

2. Sathipati SY, Ho S. Identifying a miRNA signature for predicting the stage of breast cancer. Sci Rep. 2018;8(1). doi:10.1038/s41598-018-34604-3.

3. Pezuk JA, Miller TLA, Bevilacqua JLB, Barros ACSD, Andrade F, Macedo LFA, et al. Measuring plasma levels of three microRNAs can improve the accuracy for identification of malignant breast lesions in women with BI-RADS 4 mammography. Oncotarget. 2017;8(48):83940. doi:10.18632/oncotarget.20806.

4. Liu L, Lin W, Cao S, Liu Y, Gao S, Jiao N, et al. BioDecoder: A miRNA bio-interpretable neural network model for noninvasive diagnosis of breast cancer. medRxiv. 2023 Feb 2. doi:10.1101/2023.01.31.23285308.

5. Liu X, Chang Q, Wang H, Qian H, Jiang Y. Discovery and function exploration of microRNA-155 as a molecular biomarker for early detection of breast cancer. Breast Cancer. 2021;28(4):806. doi:10.1007/s12282-021-01215-2.

6. Maurer J, Rübner M, Kuo C, Klein B, Franzen J, Wittenborn J, et al. Machine learning approach identifies miRNA signatures for breast cancer detection and classification from patient urine samples. Res Sq. 2024 Feb 29. doi:10.21203/rs.3.rs-3993094/v1.

7. Wang F, Wang J, Zhang H, Fu B, Zhang Y, Jia Q, et al. Diagnostic value of circulating miR-155 for breast cancer: a meta-analysis. Front Oncol. 2024;14. doi:10.3389/fonc.2024.1374674.

8. İmani S, Zhang X, Hosseinifard H, Fu S, Fu J. The diagnostic role of microRNA-34a in breast cancer: a systematic review and meta-analysis. Oncotarget. 2017;8(14):23177. doi:10.18632/oncotarget.15520.

9. Wang M, Gu Y, Zhang J, Jia Z, Wu H. The circulating level of non-coding RNA for breast cancer diagnosis. Res Sq. 2020 May 6. doi:10.21203/rs.2.19596/v2.

10. Koçak B, Ponsiglione A, Stanzione A, Bluethgen C, Santinha J, Ugga L, et al. Bias in artificial intelligence for medical imaging: fundamentals, detection, avoidance, mitigation, challenges, ethics, and prospects. Diagn Interv Radiol. 2024 Jul 2. doi:10.4274/dir.2024.242854.

11. Jayakumar S, Sounderajah V, Normahani P, Harling L, Markar SR, Ashrafian H, et al. What are the quality assessment standards used in artificial intelligence diagnostic accuracy systematic reviews? Res Sq. 2021 Mar 23. doi:10.21203/rs.3.rs-329433/v1.

12. Chowdhury M, Cervantes EG, Chan WY, Seitz D. Use of machine learning and artificial intelligence methods in geriatric mental health research involving electronic health record or administrative claims data: a systematic review. Front Psychiatry. 2021;12. doi:10.3389/fpsyt.2021.738466.

13. Guni A, Sounderajah V, Whiting P, Bossuyt PM, Darzi A, Ashrafian H. Revised tool for the quality assessment of diagnostic accuracy studies using AI (QUADAS-AI): protocol for a qualitative study. JMIR Res Protoc. 2024;13. doi:10.2196/58202.

14. Topor M, Pickering J, Mendes AB, Bishop D, Büttner F, Elsherif MM, et al. An integrative framework for planning and conducting non-intervention, reproducible, and open systematic reviews (NIRO-SR). 2020 Dec 14. doi:10.31222/osf.io/8gu5z.

15. Uttley L, Montgomery P. The influence of the team in conducting a systematic review. Syst Rev. 2017;6(1). doi:10.1186/s13643-017-0548-x.

16. Desmeules R, Campbell S, Dorgan M. Acknowledging librarians’ contributions to systematic review searching. J Can Health Libr Assoc. 2016;37(2). doi:10.5596/c16-014.

17. Page MJ, McKenzie JE, Bossuyt P, Boutron I, Hoffmann T, Mulrow CD, et al. The PRISMA 2020 statement: an updated guideline for reporting systematic reviews. 2021 Jan 1. doi:10.17615/pe64-en90.

18. McNutt M, Bradford M, Drazen JM, Hanson B, Howard B, Jamieson KH, et al. Transparency in authors’ contributions and responsibilities to promote integrity in scientific publication. Proc Natl Acad Sci U S A. 2018;115(11):2557. doi:10.1073/pnas.1715374115.

19. Hemmler YM, Martin F, Gezer T, Ifenthaler D. Best practices for conducting systematic reviews: perspectives of experienced systematic review researchers in educational sciences. Technol Knowl Learn. 2025;30(1):1. doi:10.1007/s10758-025-09819-9.

20. Suri H. Ethical considerations of conducting systematic reviews in educational research. 2019:41. doi:10.1007/978-3-658-27602-7_3.

21. Sawicka-Gutaj N, Gruszczyński D, Guzik P, Mostowska A, Walkowiak J. Publication ethics of human studies in the light of the Declaration of Helsinki: a mini-review. J Med Sci. 2022;91(2). doi:10.20883/medical.e700.

22. Uyisenga JP, Debit A, Poulet C, Frères P, Poncin A, Thiry J, et al. Differences in plasma microRNA content impair microRNA-based signature for breast cancer diagnosis in cohorts recruited from heterogeneous environmental sites. Sci Rep. 2021;11(1). doi:10.1038/s41598-021-91278-0.

23. Cui X, Li Z, Zhao Y, Song A, Shi Y, Hai X, et al. Breast cancer identification via modeling of peripherally circulating miRNAs. PeerJ. 2018;6. doi:10.7717/peerj.4551.

24. Zou R, Loke SY, Tan VK, Quek ST, Jagmohan P, Tang YC, et al. Development of a microRNA panel for classification of abnormal mammograms for breast cancer. Cancers (Basel). 2021;13(9):2130. doi:10.3390/cancers13092130.

25. Zou R, Loke SY, Tang YC, Too H, Zhou L, Lee ASG, et al. Development and validation of a circulating microRNA panel for the early detection of breast cancer. Br J Cancer. 2022;126(3):472. doi:10.1038/s41416-021-01593-6.

26. Gómez-Acebo I, Llorca J, Alonso-Molero J, Díaz-Martínez M, Pérez-Gómez B, Amiano P, et al. Circulating miRNAs signature on breast cancer: the MCC-Spain project. Eur J Med Res. 2023;28(1):480. doi:10.1186/s40001-023-01471-2.

27. Al-Sowayan BS, Alshareeda AT. Nanogenomics and artificial intelligence: a dynamic duo for the fight against breast cancer. Front Mol Biosci. 2021;8. doi:10.3389/fmolb.2021.651588.

28. Ma L, Gao Y, Huo Y, Tian T, Hong G, Li H. Integrated analysis of diverse cancer types reveals a breast cancer-specific serum miRNA biomarker through relative expression orderings analysis. Breast Cancer Res Treat. 2024;204(3):475. doi:10.1007/s10549-023-07208-3.

29. Jang JY, Kim YM, Kang K, Kim KS, Park Y, Kim CG. Multiple microRNAs as biomarkers for early breast cancer diagnosis. Mol Clin Oncol. 2021;14(2). doi:10.3892/mco.2020.2193.

30. Shen J, Hu Q, Schrauder M, Yan L, Wang D, Medico L, et al. Circulating miR-148b and miR-133a as biomarkers for breast cancer detection. Oncotarget. 2014;5(14):5284. doi:10.18632/oncotarget.2014.

31. Leidner RS, Li L, Thompson CL. Dampening enthusiasm for circulating microRNA in breast cancer. PLoS One. 2013;8(3). doi:10.1371/journal.pone.0057841.

32. Neagoe C, Gundershausen M, Ionică M, Neagoe OC. MicroRNAs in breast cancer: diagnostic and prognostic potential, challenges, and clinical reliability. Biomedicines. 2026;14(3):502. doi:10.3390/biomedicines14030502.

33. Giordano L, Gallo F, Petracci E, Chiorino G, Segnan N. The ANDROMEDA prospective cohort study: predictive value of combined criteria to tailor breast cancer screening and new opportunities from circulating markers: study protocol. BMC Cancer. 2017;17(1). doi:10.1186/s12885-017-3784-5.

